# Structure of a polymorphic repeat at the *CACNA1C* schizophrenia locus

**DOI:** 10.1101/2024.03.05.24303780

**Authors:** Raquel Moya, Xiaohan Wang, Richard W. Tsien, Matthew T. Maurano

**Affiliations:** Institute for Systems Genetics, NYU School of Medicine, New York, NY 10016, USA; Neuroscience Institute, NYU School of Medicine, New York, NY 10016, USA; Department of Neuroscience and Physiology, New York University, New York, NY 10016, USA; Department of Pathology, NYU School of Medicine, New York, NY 10016, USA

**Keywords:** Variable-number tandem repeat, calcium channel, schizophrenia, long-read genome assemblies

## Abstract

Genetic variation within intron 3 of the *CACNA1C* calcium channel gene is associated with schizophrenia and other neuropsychiatric disorders, but analysis of the causal variants and their effect is complicated by a nearby variable-number tandem repeat (VNTR). Here, we explored the structure and population variability of the *CACNA1C* intron 3 VNTR using 155 long-read genome assemblies from 78 diverse individuals. Based on sequence differences among repeat units, we clustered individual sequences into 7 VNTR structural alleles called Types. Three Types were related through large duplications, but the other Types diverged much earlier such that only 12 repeat units at the 5′ end of the VNTR were shared across most Types. The most diverged Types were rare and present only in individuals with African ancestry, but a multiallelic structural polymorphism was present across populations at different frequencies, consistent with expansion of the VNTR preceding the emergence of early hominins. We demonstrated that this polymorphism was in complete linkage disequilibrium with fine-mapped schizophrenia variants from genomewide association studies (GWAS), and that this risk haplotype was associated with decreased *CACNA1C* gene expression in the brain. Our work suggests that sequence variation within a human-specific VNTR affects gene expression, and provides a detailed characterization of new alleles at a flagship neuropsychiatric locus.

**SIGNIFICANCE:** Genome-wide association studies identify an association between neuropsychiatric disorders and non-coding variants within the gene *CACNA1C*, which encodes a functionally important Ca^2+^ channel in neurons. However, the variant(s) responsible for disease risk and their functional consequences are undetermined. These schizophrenia-associated SNPs are near a poorly genotyped repeat, suggesting the repeat might also contribute to disease risk. We use long-read genome assemblies to characterize the genetic diversity of this repeat, its relationship to schizophrenia-associated SNPs, and its evolutionary origins. We find that the schizophrenia risk signal is associated with reduced *CACNA1C* gene expression, and both are tightly tied to the repeat. Our analysis of repeat variation at *CACNA1C* will enable targeted investigation of regulatory mechanisms underpinning risk for schizophrenia.

## INTRODUCTION

Genome-wide association studies (GWAS) hold great promise to decipher disease biology, but their utility has been curbed by the challenges of identifying causal variants, target genes, and relevant cellular contexts (1–3). A prime example is schizophrenia, a severe and etiologically complex neuropsychiatric disorder with few effective treatments despite high heritability (70%) and incidence (1%) (4, 5). Realizing the promise of translating neuropsychiatric genetics into mechanistic insights and treatments will require systematic analysis of key association loci.

A top GWAS signal for schizophrenia (6–8) and other neuropsychiatric disorders (9–15) lies deep within the 328-kb third intron of the 645-kb *CACNA1C* gene, well away from other potential target genes (**Fig. 1A**). While the effect sizes for these schizophrenia-associated SNPs are low with odds ratios around 1.1, they are highly statistically significant. *CACNA1C* encodes the pore-forming subunit of CaV1.2, the predominant L-type voltage-gated calcium channel expressed in human central nervous system neurons (16–18). CaV1.2 channels on the somatodendritic membrane play a dominant role in triggering a signaling cascade that culminates in the phosphorylation of the nuclear transcription factor CREB (19–23), which is important for learning and memory across the evolutionary tree (24–26). A single missense variant in *CACNA1C* is responsible for the monogenic disorder Timothy Syndrome (27–29) which, along with other *CACNA1C*-related disorders (30), includes neuropsychiatric symptoms. The broader calcium channel gene family is robustly implicated in schizophrenia and other neuropsychiatric disorders (31, 32). Thus *CACNA1C* has well-established relevance for neuropsychiatric disorders, but the genetic and molecular underpinnings of the GWAS association remain unclear.

**Fig. 1.**
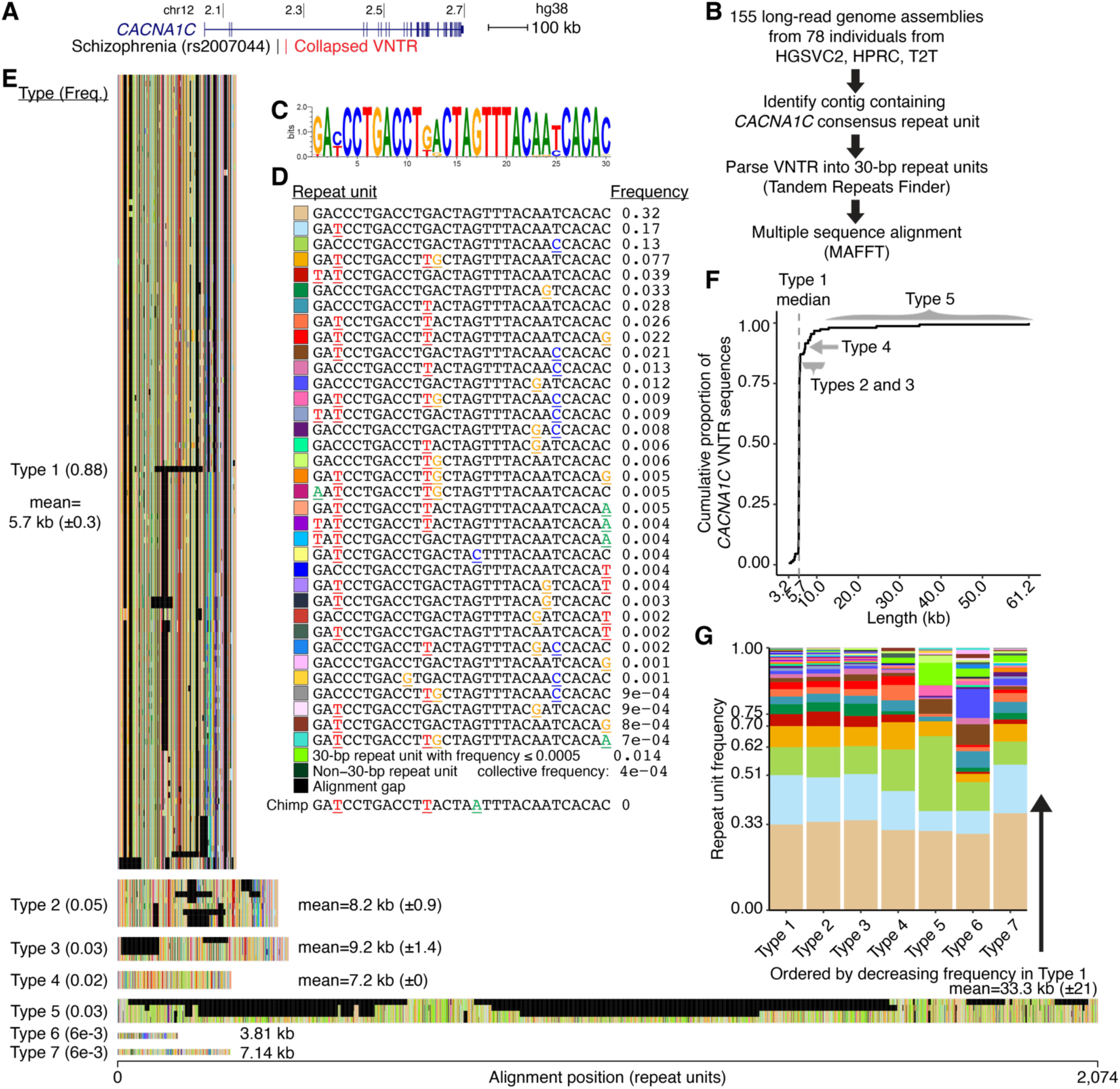
Structure of the *CACNA1C* intron 3 VNTR. (***A***) The *CACNA1C* gene overlaps a lead schizophrenia GWAS SNP (6) and the VNTR. (***B***) VNTR analysis strategy. (***C***) Sequence logo of the VNTR consensus. Letter height is proportional to frequency across all sequences. (***D***) Repeat unit color key for (***E****,**G***). 35 common repeat units (frequency > 0.0005) are shown as different colors. Nucleotide differences from the consensus repeat unit are colored and underlined. Remaining low-frequency repeat units are distinguished by length (either 30 bp or not). The chimpanzee genome contains a single copy slightly diverged from the human consensus. (***E***) Collection of 155 VNTR sequences grouped by Type and aligned. (***F***) Cumulative distribution of *CACNA1C* VNTR lengths. (***G***) Repeat unit frequencies by Type. Colors are as in (***D***). Stacked bars are ordered from bottom to top by decreasing repeat unit frequency in Type 1.

The *CACNA1C* association locus lies in close proximity to a human-specific variable number tandem repeat (VNTR) (33). Tandem repeats have been implicated in a wide variety of human traits and diseases (34), including Amyotrophic Lateral Sclerosis (ALS) (35), Alzheimer’s disease (36), progressive myoclonic epilepsy (37), and others. Tandem repeats show widespread association with expression of nearby genes (38–42), and can modulate transcription factor binding (43). Complex variants like VNTRs can be tagged by surrounding SNPs (44–48); for example at the *C4* schizophrenia locus where GWAS SNPs are linked to a structural variant that affects gene expression (48). Thus several lines of evidence suggest the involvement of VNTRs in the regulation of gene expression and disease.

However, typical genomic analyses exclude repetitive regions. VNTRs, defined by repeat units longer than 6 bp, are especially complex (49) and difficult to analyze. Indeed, the *CACNA1C* VNTR is collapsed to 300 bp in the human reference genome (33). Thus, testing whether the *CACNA1C* schizophrenia association is related to this repeat requires a systematic definition of VNTR variants. Specialized tools can estimate VNTR length from short-read data (38, 39, 50), but accurate inference of repeat structure and variation is difficult when VNTRs exceed sequencing read lengths. The recent development of high-quality long-read assemblies offers an opportunity for detailed assessment of polymorphic VNTR sequences (50–54) which can facilitate their assessment in disease studies.

Here we analyzed the *CACNA1C* VNTR in 155 long-read haplotype assemblies and characterized three aspects of its genetic variation: (*i*) a polymorphic structure and its likely basis in duplication, (*ii*) linkage disequilibrium between common *CACNA1C* VNTR polymorphism and schizophrenia-associated SNPs, and consequences for *CACNA1C* gene expression, and (*iii*) ancestries of *CACNA1C* VNTR polymorphism that suggest its evolutionary history. Our analysis uncovers unexpected patterns of structural diversity within the repeat and delineates a series of common alleles at two variable regions within the predominant VNTR sequence type. We show that one of these variable regions is tightly correlated with schizophrenia risk and reduced *CACNA1C* gene expression in brain tissue. Our work thus serves as an example for investigation of other difficult association loci.

## RESULTS

### Structural characterization of the *CACNA1C* VNTR in long-read assemblies

To provide a foundation for future study of its potential role in disease, we started by comprehensively mapping the repeat structure and variation of the *CACNA1C* intron 3 VNTR. We analyzed phased long-read genome assemblies from the Human Genome Structural Variation Consortium Phase 2 (HGSVC2) (52), the Human Pangenome Reference Consortium (HPRC) (54), and the Telomere-to-Telomere project (53). This collection contained 155 haplotypes representing 78 distinct individuals, including 3 trios and a haploid genome assembly from the homozygous hydatidiform mole CHM13 (**Fig. 1B**, ***SI Appendix***, **Dataset S1, Table S1**). For each assembly, we identified the contig containing the previously identified consensus repeat unit of this VNTR (33) and extracted the full repeat sequence for analysis (***SI Appendix***, **Dataset S2**, **Dataset S3**). We confirmed that each identified VNTR sequence was present on a single contig and flanked by unique sequence.

Analysis of repeat units within these sequences recapitulated the known motif, with strict conservation at 22 out of 30 positions (**Fig. 1C**). We identified 34,172 repeat units in total and 158 distinct repeat units, most of which were 30 bp in length (**Fig. 1D**, ***SI Appendix***, **Dataset S4**). A subset of 35 distinct repeat units with frequency >5x10^-4^ comprised >98% of *CACNA1C* VNTR sequences. The most common repeat unit, defined as the consensus, differed from other units by an average of 2.3 nucleotide substitutions and comprised 32% of repeat units in all *CACNA1C* VNTR sequences. Of the 8 most variable nucleotide positions, 6 had only a single alternate nucleotide. Infrequent repeat units fell into two categories: repeat units of size other than 30 bp (n = 11) and infrequent 30-bp units (frequency ≤ 5x10^-4^, n = 111). Thus, while the *CACNA1C* VNTR has a strong consensus sequence, its significant variability permits structural analysis within the repeat.

To characterize the alleles of the *CACNA1C* VNTR, we iteratively performed multiple sequence alignment starting from all 155 repeat sequences. We manually grouped sequences until no sequence appeared visually misplaced, converging on 7 distinct Types of the *CACNA1C* VNTR (**Fig. 1E**). Within each Type, sequence alignments showed high consistency across nearly all positions (***SI Appendix***, **Fig. S1A**). To further assess our Type definitions in an unsupervised manner, we clustered all 155 repeat sequences. Types 1, 4, and 5 clustered separately from each other (***SI Appendix***, **Fig. S1B**). Types 2 and 3 were interspersed with Type 1 sequences, reflecting similarities in their sequences.

Each VNTR Type was supported by at least one PacBio HiFi assembly (***SI Appendix***, **Table S2**). We verified that a subset of 8 assemblies representative of each Type had uniform coverage depth of PacBio HiFi reads over the VNTR (***SI Appendix***, **Fig. S2**, **Dataset S5**). Then we generated a consensus sequence for each Type from the most frequent repeat unit at each position that best represented its distinct length and repeat unit order (***SI Appendix***, **Fig. S1A**, **Dataset S6**). Type 1 represented 88% of sequences (**Fig. 1E**) and defined the most common length (median = 5.7 kb, **Fig. 1F**). Type 2 and Type 3 sequences were 30-40% longer than Type 1 sequences (median, Type 2: 8.3 kb, Type 3: 8.4 kb) with less consistency in length. Type 4 sequences had identical lengths (7.2 kb) and very few sequence differences among them. Type 5 sequences were the longest (13-61 kb) and most variable (s.d. = 21 kb). Types 6 and 7 were defined by single sequences that were distinct from other Types in length and repeat unit order.

In each Type, at least 85% of the overall sequence was comprised of a few distinct units (n = 13 out of 158 distinct units) (**Fig. 1G**). Repeat unit frequencies were nearly identical across the tightly related Types 1, 2, and 3, while Types 4-7 were characterized by increased inclusion of rare repeat units. The most common repeat unit (shown as beige) had a similar representation within each Type, while the representation of other repeat units varied across Types. Thus a small set of repeat units prolifically recur and stereotypically compose the *CACNA1C* VNTR, with varying proportions across Types.

These Types represented dissimilar structural *CACNA1C* VNTR alleles. Only the first repeat unit was shared among all Types, though the first 12 repeat units were shared among most Types (***SI Appendix***, **Fig. S3A**). The 3′ end was also variable and no position was constant across all Types (***SI Appendix***, **Fig. S3B**). Overall, these sequences attest to an exceptionally rich diversity in repeat structure and raise questions about its mutational origins.

### Pervasive tandem duplication within the *CACNA1C* VNTR

To investigate the sequence origin of 7 Types and the source of their length variability, we scanned each Type for duplicated and unique sequence. Given the high degree of similarity between repeat unit frequencies of Types 1, 2, and 3, we reasoned that they may be structurally related. We computed counts of exact matches to a sliding window (width = 6 repeat units) across Type 1 for Types 1, 2, and 3 using their consensus sequences. This duplication assay identified two separate kilobase-sized tandem duplications (Duplication 1 and Duplication 2) that distinguished Type 2 and Type 3 from Type 1 (**Fig. 2A** and **B**). Duplication 1 breakpoints varied across Type 2 sequences (**Fig. 1E**, ***SI Appendix***, **Fig. S4**), while Duplication 2 breakpoints in Type 3 sequences were identical (**Fig. 1E**). We identified a single Type 3 sequence (NA21309_paternal) that had a third copy of Duplication 2 with these breakpoints (**Fig. 2B**), indicating this site could be prone to instability. These results suggest that Type 2 and Type 3 are derived from Type 1.

**Fig. 2.**
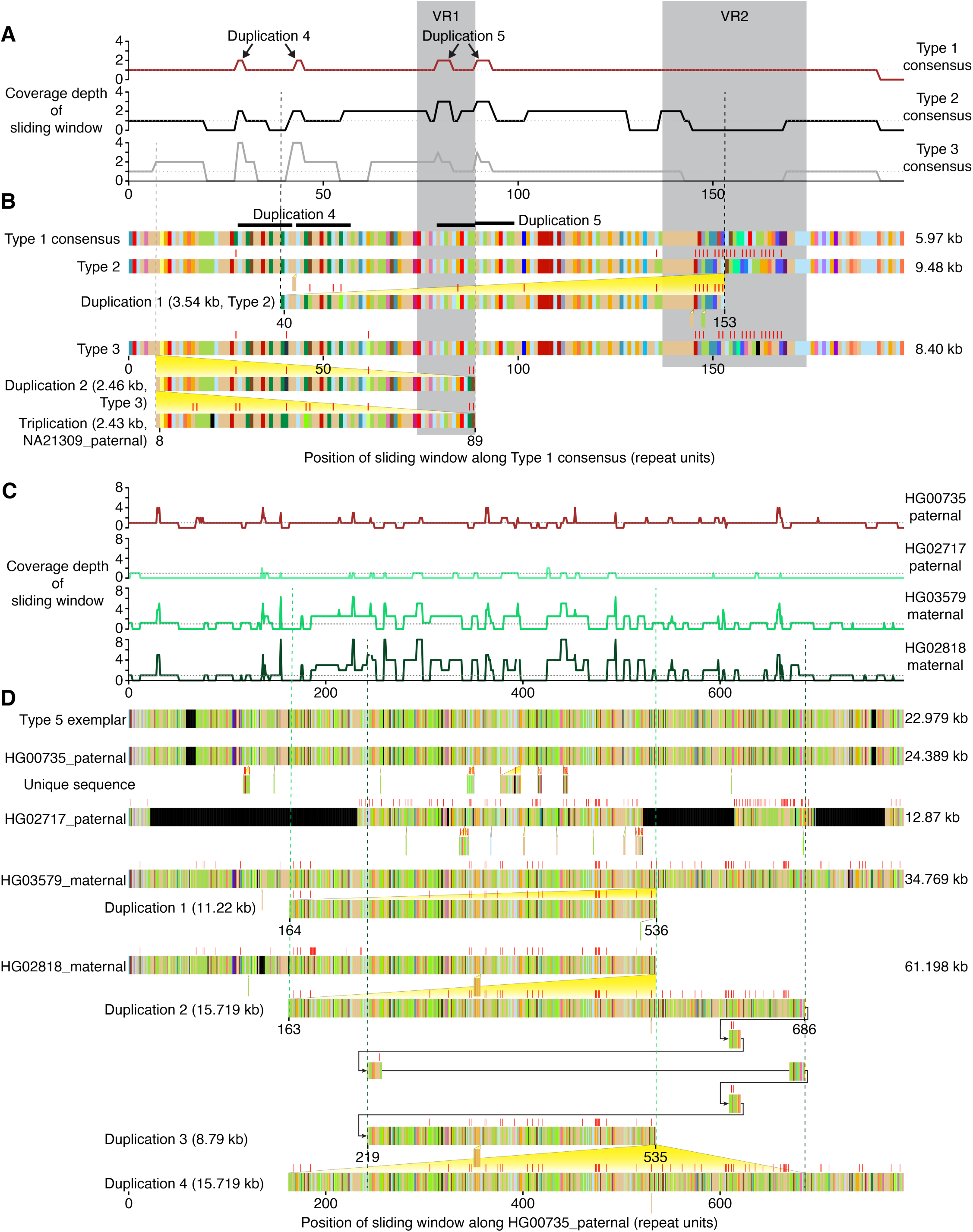
Large duplications within *CACNA1C* VNTR sequences. Analysis of Types 2 and 3 (***A*** and ***B***), and Type 5 (***C*** and ***D***) VNTR sequences. (***A*** and ***C***) Y-axis measures coverage depth of a sliding window across an exemplar sequence (x-axis). Sliding window width is 6 (***A***) or 8 (***C***) repeat units. Type 1 consensus was used as the exemplar in (***A***). For (***C***), a Type 5 exemplar was derived from HG00735_paternal by removing unique sequence not found in other Type 5 sequences. Large tandem duplications are indicated by vertical dashed lines (paired by color) and yellow triangles. Arrows in (***D***) indicate tiling paths. Repeat unit colors are as in Fig. 1D. Non-gap mismatches to the exemplar sequence are shown with red ticks.

Two smaller tandem duplications were identified in Types 1, 2, and 3 using this approach. A 420-bp duplication (Duplication 4) was not exactly in tandem, but instead was separated by one repeat unit. A 300-bp duplication (Duplication 5) overlapped part of VR1. Their partial overlap with Duplication 1 and Duplication 2 in Type 2 and Type 3 suggested that the smaller tandem duplications preceded larger ones in time. This pattern shows that one mechanism of expansion for this VNTR is successive tandem duplication of increasingly long VNTR segments.

The alignment of Type 5 sequences showed significant heterogeneity (**Fig. 1E**). Scanning for duplications within Type 5 revealed multiple kilobase-sized tandem duplicated segments (**Fig. 2C**). To facilitate analysis of Type 5, we established a Type 5 exemplar sequence from HG00735_paternal, omitting several small insertions not found in other Type 5 sequences (**Fig. 2D**, **Fig. 1E**). HG00735_paternal was selected because it had a single exact match to the sliding window across most of the sequence. The other Type 5 sequences showed structural variety relative to this sequence: HG02717_paternal contained large deletions, HG03579_maternal had a kilobase-sized Duplication 1, and HG02818_maternal contained a complex tiling path starting with Duplication 2 and proceeding with multiple smaller interspersed duplications, followed by two more kilobase-scale duplications, suggesting genesis of this sequence through a complex pattern of expansion (**Fig. 2D**). This degree of structural variation suggests that Type 5 may be particularly prone to rearrangement.

These results indicate that the increased length of Type 2, 3, and 5, as well as heterogeneity within each Type, is governed by kilobase-scale duplication. We also scanned for duplications within the Type 4, 6, and 7 consensus sequences, and did not find large duplications, but did observe a handful of smaller tandem duplications (***SI Appendix***, **Fig. S5**). We next sought to determine if Type 1 also harbored evidence of duplication. We scanned the alignment of Type 1 sequences to identify regions of shared and variable sequence by computing a positional variability score (**Fig. 3A**). The least variable parts of the sequence overlapped 26 alignment gaps (**Fig. 3B**), most frequently 30 bp in length but ranging up to 300 bp (***SI Appendix***, **Fig. S6A**, **Dataset S7**). We analyzed the sequences overlapping each gap and found that 24 out of 26 gaps corresponded to tandem duplications (**Fig. 3C**). Most of the duplicated sequences occurred without divergence (***SI Appendix***, **Fig. S6B**). These duplications were rare individual events (***SI Appendix***, **Fig. S6C**), but collectively common (n = 102 duplications total, 0.76 duplications per Type 1 sequence). Two alignment gaps near the end of the *CACNA1C* VNTR overlapped the most variable region of the sequence and were inconsistent with this model of tandem duplication (**Fig. 3C**, red arrows).

**Fig. 3.**
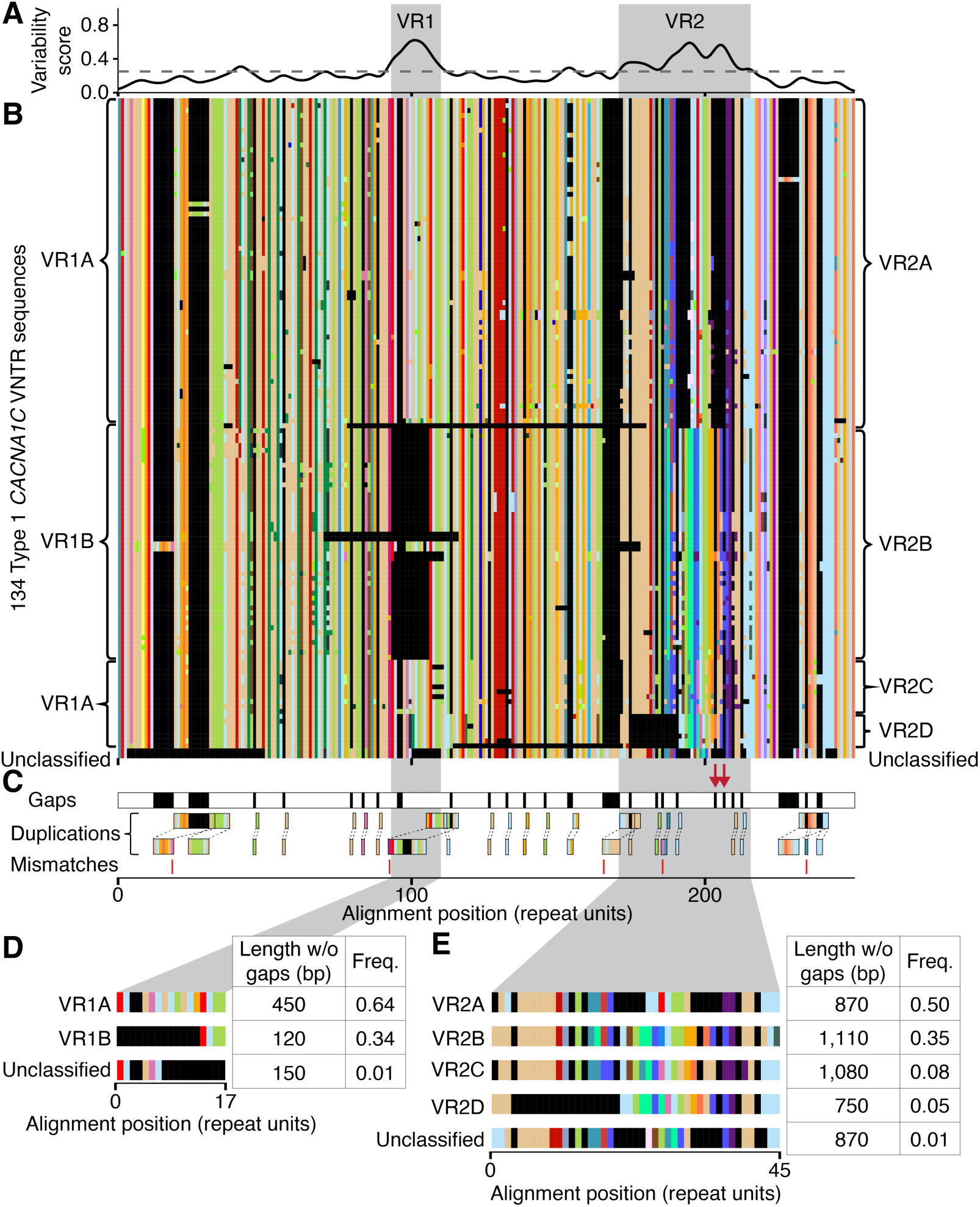
Characterization of tandem duplications and Variable Region alleles in Type 1 sequences. (***A***) Repeat unit variability is shown as normalized Shannon’s uncertainty H(x). Variable regions are defined as H(x) > 0.25 and length > 7 aligned repeat units. (***B***) Multiple sequence alignment of Type 1 sequences. Variable region boundaries are labeled with gray boxes. (***C***) Analysis of 26 alignment gaps in the Type 1 consensus sequence. 24 out of 26 gaps correspond to tandem duplications. The most common duplication overlapping each gap is shown. Five gaps overlap multi-allelic duplications (***SI Appendix***, **Dataset S7**). Two gaps in Variable Region 2 are not consistent with this model (red arrows). Mismatches (in nucleotides) between duplication source and destination sequences are shown with red ticks. (***D*** and ***E***) Variable Region 1 and 2 (VR1 and VR2) alleles, their unaligned lengths (in bp), and their frequencies in 134 Type 1 sequences. “Unclassified” includes alleles that do not cluster near the consensus (***SI Appendix***, **Fig. S8A**, **Fig. S9A**) and whose full VNTR sequences have a large aligned edit distance to exemplar *CACNA1C* VNTR sequences (***SI Appendix***, **Fig. S8B**, **Fig. S9B**). (***B-E***) Repeat unit colors defined in Fig. 1D.

Thus the *CACNA1C* VNTR shows pervasive tandem duplication (large and small), whereas a second mutational process may be at play over its most variable region. Rare Types 4, 6, and 7 diverged such that the extant sequences in this dataset show no evidence of their history, yet we speculate they arose through tandem duplication too. In contrast, Types 1-3 represent the most common structure and collectively account for 95% of sequences, thus we sought to examine common sequence variation within this structure that may be linked to the GWAS association signal.

### Common *CACNA1C* repeat polymorphisms

Within Type 1 sequences, two previously identified regions (33) stood out because of their high variability: Variable Region 1 (VR1) and Variable Region 2 (VR2) (**Fig. 3A** and **B**).We sought to parse variation at VR1 and VR2 into several groups of related sequences that delineate multiple alleles at each VR (***SI Appendix***, **Fig. S7A**). We tabulated 18 distinct VR1 sequences spanning 17 aligned repeat units. We grouped them into two alleles and identified consensus sequences for VR1A and VR1B (**Fig. 3D**, ***SI Appendix***, **Fig. S8**, **Dataset S8**). VR1A and VR1B differed by 11 repeat units and 330 positions at the nucleotide level. VR1B lacked a 330-bp segment found in VR1A, suggesting that these alleles resulted from ancestral expansion or truncation of each other. VR1A had a frequency of 64%, higher than VR1B (***SI Appendix***, **Fig. S7B**). Most VR1 sequences matched the consensus sequence and any VR1 sequence that differed from it had an average of 2 repeat units changed. One VR1 sequence found in two individuals was left unclassified due to the high number of mismatches to either VR1A or VR1B.

For VR2, we tabulated 71 distinct VR2 sequences spanning 45 aligned repeat units. We grouped VR2 sequences into four alleles: VR2A, VR2B, VR2C, and VR2D (**Fig. 3E**, ***SI Appendix***, **Fig. S9**, **Dataset S9**). The large number of repeat unit differences between VR2 alleles corresponded to a large number of differences at the nucleotide level (including alignment gaps) (***SI Appendix***, **Fig. S9B**, **Table S3**). Only 25% of sequences matched a VR2 allele sequence exactly, but all sequences had fewer than 25% of repeat units deviating from the consensus, which corresponded to a low number of intra-allele nucleotide differences (***SI Appendix***, **Table S4**). One VR2 sequence was unclassified due to the high number of mismatches to any other VR2 allele. The four VR2 alleles formed two common alleles and two rarer alleles (***SI Appendix***, **Fig. S7C**) with similar lengths. These alleles were defined by different repeat units and their orders (**Fig. 3E**), suggesting that VR2 derived from a more complex mutational process than VR1.

Comparison of two full *CACNA1C* VNTR sequences with different VR1 and VR2 alleles confirmed that VR1 and VR2 alleles corresponded to distinct sequences at the nucleotide level (***SI Appendix***, **Fig. S8B**, **Fig. S9B**). Ordering Type 1 sequences by VR2 allele revealed high LD between VR1 and VR2 (R^2^ = 0.85, **Table S5**): VR1A was found in the same sequences as VR2A, VR2C, and VR2D; VR1B was found in the same sequences as VR2B (***SI Appendix***, **Fig. S7**). The unclassified VR1 and VR2 sequences co-occurred in the same assemblies. VR1 and VR2 alleles were significantly expanded compared to the previously published description (33) (***SI Appendix***, **Fig. S10**). Thus, the most common Type 1 sequences include two variable regions in high LD, each having two common alleles that might play a key role in functional genetic variation.

### Variable Region 2 is associated with schizophrenia and *CACNA1C* expression

Intron 3 of *CACNA1C* harbored a series of DNaseI hypersensitive sites (DHSs) in fetal brain from the Roadmap Epigenomics Mapping Consortium (1). These DHSs were also active across a variety of neuronal subtypes including glutamatergic neurons from fetal brain, as shown by scATAC-seq (55). The VNTR lay among *CACNA1C* eQTLs in four brain tissues profiled by the Genotype-Tissue Expression (GTEx) project (56): cerebellum (n = 130), cerebellar hemisphere (n = 130), with 122 eQTLs shared between these replicate tissues, putamen (n = 1), and substantia nigra (n = 1) (**Fig. 4A**).

**Fig. 4.**
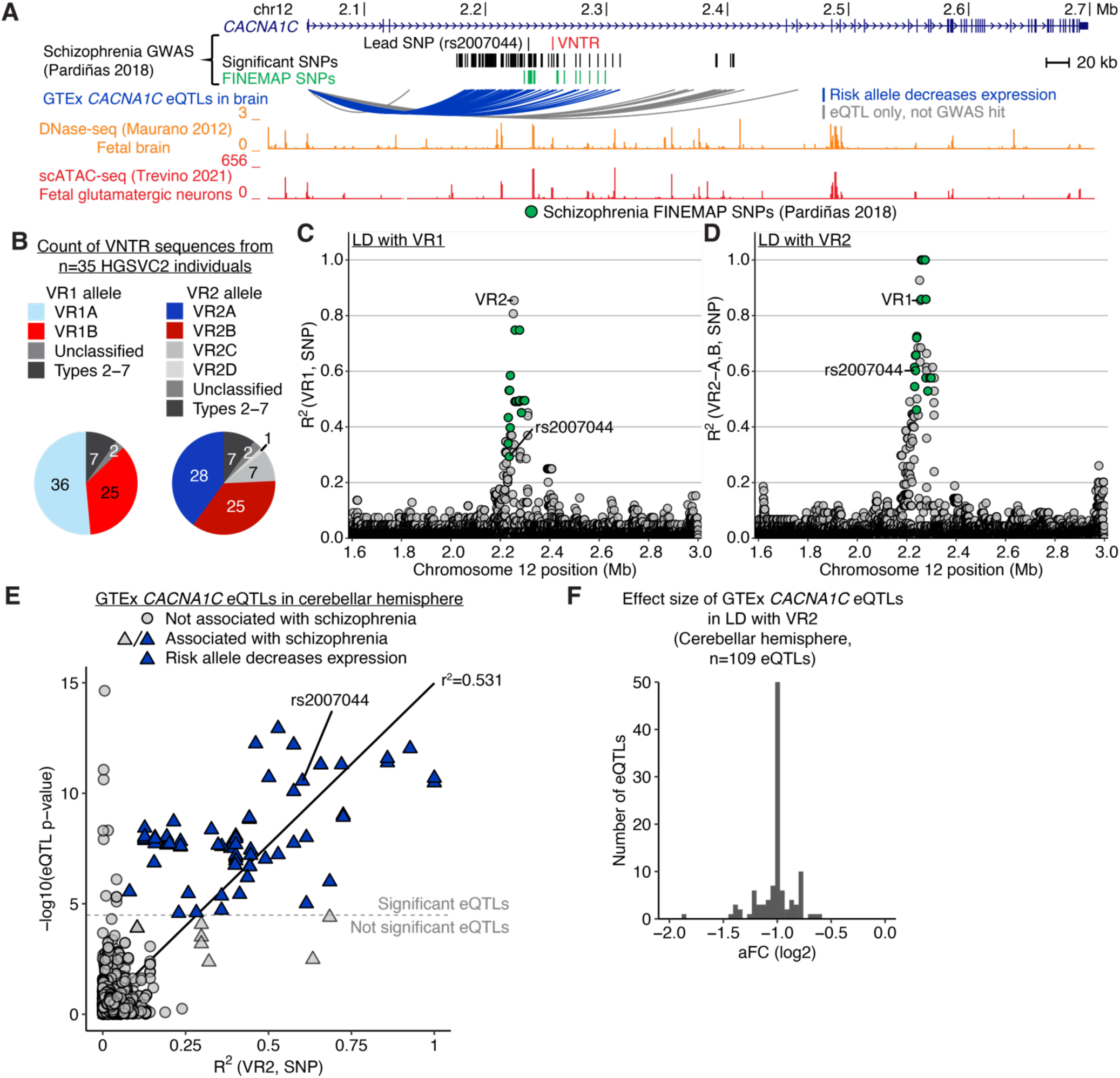
Schizophrenia risk haplotype decreases *CACNA1C* expression and is in linkage disequilibrium with Variable Region 2. (***A***) *CACNA1C* locus showing schizophrenia-associated SNPs (P < 5x10^−8^), the fine-mapped causal set (PP > 95%) (6), and the VNTR (33). Arcs depict GTEx brain eQTLs (cerebellum, cerebellar hemisphere, putamen, and substantia nigra). Shown are DNaseI hypersensitive sites (DHSs) in fetal brain from the Roadmap Epigenomics Mapping Consortium (1) and scATAC-seq data in in glutamatergic neurons from fetal brain (55). Multi-mapping DNase-seq reads are included. (***B***) VR allele counts for 35 HGSVC2 individuals with phased SNP genotypes. Alleles shown in gray were excluded from linkage disequilibrium (LD) analysis. (***C*** and ***D***) LD (y-axis) between VR1 (***C***) or VR2 (***D***) and surrounding SNPs. Fine-mapped schizophrenia SNPs are highlighted in green. LD was calculated for VR2A and VR2B alleles; LD with VR2C and VR2D is shown in ***SI Appendix***, **Fig. S11**. Three fine-mapped schizophrenia SNPs are in perfect LD with VR2 (partially overlapping points at R^2^=1.0). (***E***) Analysis of the relationship between VR2 and eQTLs in cerebellar hemisphere. eQTL significance in brain (y-axis) is strongly correlated with VR2 LD (x-axis). Of the eQTLs associated with schizophrenia (triangles), those in high LD with VR2 (blue triangles) all show decreased expression of *CACNA1C*. Dashed gray line indicates *P* value threshold for cerebellar hemisphere eQTLs. (***F***) Histogram of *CACNA1C* effect sizes for cerebellar hemisphere eQTLs in LD with schizophrenia and VR2 (blue triangles from ***E***).

To fine-map VR1 and VR2 among the schizophrenia association at *CACNA1C*, we first examined linkage disequilibrium (LD) between VR1, VR2, and the nearby schizophrenia-associated SNPs (6). We focused on a subset of long-read assemblies with available phased SNP genotypes (n = 70 HGSVC2 haplotypes) (52). This subset of *CACNA1C* VNTR sequences had VR allele frequencies comparable to the whole dataset (**Fig. 4B**, ***SI Appendix***, **Fig. S7B** and **C**). We calculated LD between each VR and SNPs in a 1.4 Mb region around *CACNA1C* (**Fig. 4C** and **D**, ***SI Appendix***, **Fig. S11**). Fine-mapped schizophrenia SNPs showed high LD with VR2 and moderate LD with VR1, while SNPs not associated with disease were less correlated with both. Three fine-mapped schizophrenia SNPs were in perfect LD with VR2. We calculated LD in the subset of European HGSVC2 individuals (n = 7) whose ancestry is most similar to the original GWAS cohort (6) and found that the LD pattern of these individuals is similar to the full HGSVC2 dataset (***SI Appendix***, **Fig. S12**). Including Types 1, 2, and 3 in an analysis of LD between SNPs and VRs showed similar patterns as Type 1 sequences alone (***SI Appendix***, **Fig. S13**, **Fig. S14**).

We used these data to examine the relationship between *CACNA1C* gene expression, the schizophrenia association, and VR2. Intersecting *CACNA1C* eQTLs with GWAS results showed that schizophrenia risk alleles were associated with reduced *CACNA1C* expression. Comparison of eQTL *P* values to LD with VR2 showed that the eQTL was composed of two distinct eQTL signals (**Fig. 4E**, ***SI Appendix***, **Fig. S15**): one signal was associated with schizophrenia and VR2; the other was not associated with either schizophrenia or VR2. The degree of LD with VR2 was strongly correlated with the statistical significance of *CACNA1C* eQTLs (R^2^ = 0.531, P < 2.2x10^−16^). eQTL effect sizes were quantified as log allelic fold change (aFC), which is equivalent to the log-fold expression ratio of the individuals homozygous for the alternate allele to those homozygous for the reference allele of an eQTL (57). Cerebellar hemisphere eQTLs in LD with VR2 had an average aFC of –0.998, reflecting a two-fold lower expression than protective alleles (**Fig. 4F**). Thus, our analysis identifies VR2 as a potentially functional variant at this locus.

### Ancestry and history of the *CACNA1C* VNTR

The individuals in our dataset included a mixture of African, Asian, American, and European ancestries (**Fig. 5A**). Type 1 sequences reflected this overall composition, while Types 2 and 3 showed an increased representation of African and Asian ancestries, respectively, suggesting two separate divergence events that are now segregated geographically (**Fig. 5B**). Types 4, 5, 6, and 7 were found mostly in African individuals. The ancestry of Type 1 can be further broken down by VR2 allele, where African ancestry showed a higher prevalence of VR2B and VR2D, East Asian ancestry showed a higher prevalence of VR2C, and South Asian ancestry showed a higher prevalence of VR2B (**Fig. 5C**). The two unclassified VR2 sequences were exclusively African, consistent with high genetic diversity rather than sequencing errors. The ubiquity of Type 1 suggests it may be the ancestral allele relative to the other Types.

**Fig. 5.**
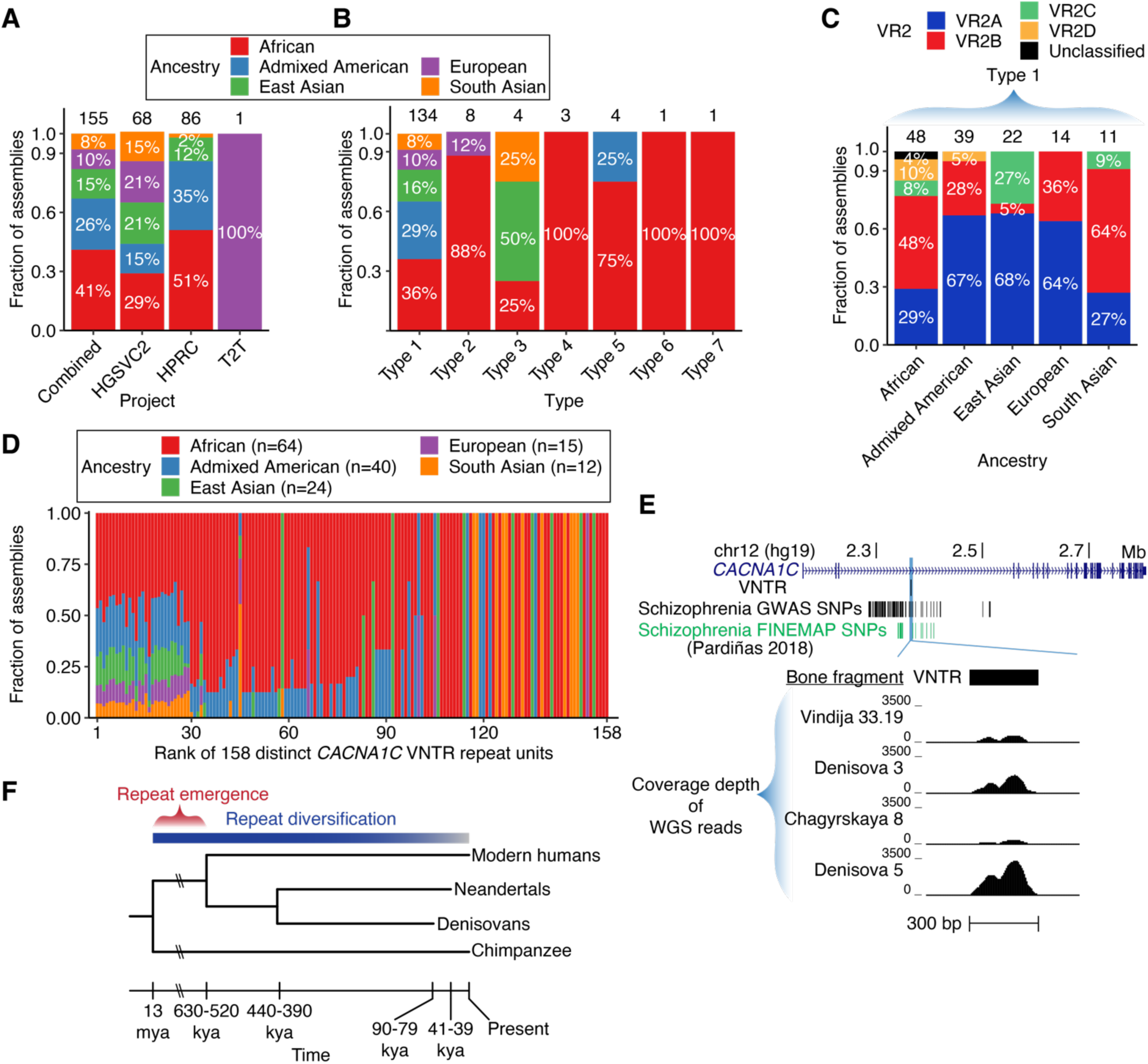
Evolutionary history of *CACNA1C* VNTR diversity. (***A*** and ***B***) Ancestry frequency in 155 haplotype assemblies (***A***) and in seven VNTR Types (***B***). Numbers above indicate count of haplotype assemblies. (***C***) Abundance of each VR2 allele by ancestry. Ancestry of each individual is defined by its 1000 Genomes super-population. (***D***) Fraction of assemblies by ancestry for each distinct repeat unit. A haplotype assembly is included if its VNTR sequence contains at least 1 copy of the repeat unit. The 158 repeat units (x-axis) are ordered by decreasing frequency and numbered by rank. (***E***) Pileup of reads over the *CACNA1C* VNTR region (GRCh37/hg19) from whole-genome sequencing of four ancient hominin genomes. Tracks are ordered from most recent to most historic date that the hominin individual lived (***SI Appendix***, **Table S6**, top to bottom). (***F***) Schematic of primate evolutionary tree with time range of VNTR emergence. References for the date of each event are in ***SI Appendix***, **Dataset S10** (62, 87).

The fraction of assemblies by ancestry containing each distinct *CACNA1C* VNTR repeat unit varied by repeat unit frequency. The 29 most abundant repeat units matched the ancestry distribution of the dataset (**Fig. 5D**). The remaining 129 repeat units (frequency < 0.0011) were found either in a few sequences of predominantly African ancestry or single *CACNA1C* VNTR sequences (n = 25) of various Types (***SI Appendix***, **Dataset S4**). Taken together, our results align with the findings that the highest amount of genetic diversity, including that at structural variants (58, 59), exists within humans of African descent (60).

The *CACNA1C* VNTR has been reported to be human-specific, existing as a single 30-bp sequence in chimpanzee and other non-human primates (33). Chimpanzees and gorillas have matching single 30-bp sequences that differ from the human *CACNA1C* VNTR consensus repeat unit at three nucleotides (**Fig. 1D**) (33). The Ts at the 3rd and 12th positions of the 30-bp chimpanzee sequence were observed in 42% and 21% of human repeat units, respectively. In contrast, the A at the 17th position of the 30-bp chimpanzee sequence was never observed in 34,172 human repeat units from this dataset. The G at this position of the human consensus repeat unit is rarely variable; it is a C in only 0.44% of repeat units. Thus, by virtue of the 17th position, all known human *CACNA1C* VNTR repeat units are distinct from the sequence in our closest primate relative.

To investigate the timing of the repeat formation in ancient human populations, we examined four ancient genomes dating to approximately 52-122 thousand years ago: three Neandertals (61–63) and one Denisovan (64) (***SI Appendix***, **Table S6**). We analyzed DNA sequencing coverage depth over the *CACNA1C* VNTR to infer copy number. By assuming that the coverage depth ratio over the repeat and its flanks resembles modern samples, our analysis indicated the presence of a repeat at lengths ranging from 3,360 kb to 10,560 kb, which is similar to that found in modern humans (**Fig. 5E**, ***SI Appendix***, **Table S6**, **Fig. S16**). This analysis dates the expansion to somewhere between 13 million years ago (the estimate for divergence of modern humans and chimpanzees) (62) and 630-520 thousand years ago (the estimate for divergence of modern humans and Neandertals) (62). Our results show that the *CACNA1C* VNTR had already expanded prior to the divergence of early hominin populations (**Fig. 5F**).

## DISCUSSION

Leveraging the availability of phased long-read genomes, we describe the structure and genetic diversity of a human-specific VNTR at the *CACNA1C* schizophrenia locus. We delineated the complex variation of the VNTR in terms of repeat unit variants, Types, internal duplications, and polymorphic variable regions. The rarest repeat units and longest VNTR sequences were found in Types 4-7 and depleted in individuals with European ancestry, whereas duplications ranging from a single repeat unit to more than ten kilobases were found across Types. Variable Regions within the most common VNTR structure defined distinct alleles and were found across populations. Our analysis goes beyond the 27 previously reported sequences (33), all of which are classified as Type 1 in our analysis. We showed that the schizophrenia association and *CACNA1C* brain eQTL are both tightly linked to VNTR variants. These data support a model where common variants mediate schizophrenia risk through an effect on *CACNA1C* expression. While our analysis most specifically implicates VR2, further investigation will be needed to determine the exact function of the allelic diversity at the VNTR. In particular, while our results show an association between VR2 and gene expression, it is unclear whether the VNTR acts as an enhancer at its endogenous locus, and if so, which repeat sequence features are necessary and sufficient. In particular, VR1 and VR2 lie only 1.44 kb apart on average, are in tight linkage disequilibrium, and the VNTR is surrounded by numerous intronic DNaseI hypersensitive sites which may regulate its function.

A key question is whether schizophrenia-associated variants increase or decrease CaV1.2 channel function, and in what cell context. Our results indicate that the risk allele decreases *CACNA1C* expression in cerebellum. Corroborating this, risk alleles in *CACNA1C* intron 3 have been previously associated with decreased *CACNA1C* expression in cerebellum (65) and superior temporal gyrus (66). Assessment of repeat-associated k-mers in GTEx is also consistent with the risk allele reducing *CACNA1C* expression in cerebellum (41). In contrast, studies in dorsolateral prefrontal cortex (67) and induced neurons (68) have reported an association of risk alleles with increased *CACNA1C* expression. Timothy Syndrome mutations result in a gain of CaV1.2 channel function (27, 29, 69). Examples of *CACNA1C* loss of function also exist and have neurodevelopmental and neurobehavioral sequelae (70–74). Yet, it is unclear what the specific consequences of decreased *CACNA1C* expression would be on the affected cell types and brain networks. Furthermore, schizophrenia is not thought to involve brain networks in cerebellum; instead, several cortical networks are implicated, many of which involve prefrontal brain regions (31). The high cell type uniformity of cerebellum could favor detection of *CACNA1C* eQTLs, as the cerebellum contains 80% of the neurons in the human brain, of which 99% are granule cell neurons (75, 76). Other regions including neocortex manifest greater neuronal heterogeneity and a higher prevalence of glial cells (77) that have low *CACNA1C* expression (78), making them less favorable for eQTL detection.

Repetitive sequences pose increased risk for sequencing and assembly errors. The error rates of the source assemblies have been reported to be <10^−4^ for PacBio continuous long read assemblies (HGSVC2) (52), <10^−5^ for PacBio HiFi assemblies (HGSVC2, HPRC) (52, 54), and <10^−7^ for the CHM13 haploid assembly (53). Our analyses of Types and VR alleles is tolerant to nucleotide variation as each group is supported by multiple independent sequences, with the exception of Types 6 and 7 that were represented only by a single sequence. Each genome in our analysis included exactly two VNTR sequences, each on a single contig and flanked by unique sequence. For the 5 individuals included in both HGSVC2 and HPRC, 4 had identical *CACNA1C* VNTR sequences (**Materials and Methods**). Additionally, for each of the 3 trios in this dataset, we observed accurate transmission of a single maternal and paternal *CACNA1C* VNTR sequence. No individual had two identical alleles, lowering the chance that a read could be assigned to the incorrect haplotype. Together, these considerations suggest that the general features of our VNTR analysis do not reflect errors of basecalling or assembly.

Our analysis discretizes *CACNA1C* VNTR VR sequences that exist among a spectrum of variation, providing a convenient model of complex genomic data but potentially drawing arbitrary distinctions. In particular, partitioning VR2 sequences into 4 alleles masks significant underlying variability. First, some sequences resembled both VR2B and VR2C, and their assignment to VR2C may reflect arbitrary features of our multiple sequence alignment. Second, VR2B intra-allele sequence variability was higher than the inter-allele differences between VR2B and VR2C, suggesting that VR2B and VR2C could be more similar than different. Indeed, VR2C showed high LD with schizophrenia FINEMAP SNPs (***SI Appendix***, **Fig. S11A**). But one clear difference between VR2B and VR2C is that VR2B co-occurs with VR1B, and VR2C with VR1A. This analysis underscores the question of whether variable structural alleles in high LD should be genotyped separately or together as a feature of the whole structural variant, which should be carefully considered in future analyses of VR1 and VR2.

The striking structural diversity of the *CACNA1C* VNTR and the prevalence of tandem duplication within invites speculation around its mechanism of expansion. In contrast with somatic variation at DNA repeats induced by replication slippage or homologous recombination after erroneous DNA repair, variation within subtelomeric VNTRs may emerge through recombination events; *CACNA1C* itself lies only 2 Mb from the start of chromosome 12. One possible mechanism for *CACNA1C* VNTR expansion is unequal exchange through homologous pairing-dependent events, which could change repeat unit copy number and repeat unit sequences but preserve the repeating frame (79, 80). Consistent with this, we observe only 11 distinct repeat units of size other than 30 bp; most (n = 8) are only 1 bp longer or shorter than a typical 30 bp repeat unit. Models of this mutational process show that over time such repetitive sequences would be unstable in length. Unequal crossover events between sister chromatids that lengthen or shorten the repetitive sequence may inevitably occur (79, 80). In line with this model, we observe that the most diverged Types (Types 4, 5, 6, and 7, n = 9 individuals) have Type 1 or 2 on the other allele of an individual, suggesting that changes in VNTR length could happen within a haplotype and not between two alleles. Subtelomeres demonstrate higher meiotic recombination rates and higher frequency of double-strand breaks. Coincident with this, VNTRs are enriched relative to short tandem repeats (STRs) near chromosome ends (58, 81, 82). Our data support the hypothesis that evolutionary adaptation, involving highly variable gene families, may be privileged at subtelomeric chromosome ends. Examples of subtelomeric gene families driving adaptation to environmental changes exist in human (e.g., the olfactory receptor gene family) (83), yeast (e.g., MAL gene family) (84), and the parasite causing malaria in humans *P. falciparum* (e.g., antigen genes) (85). Furthermore, the number of distinct structural alleles of the *CACNA1C* VNTR provides hints about its patterns of divergence. Our analysis suggests that Type 2 and Type 3 derive from Type 1, while the other Types that only share starting repeat units emerged more independently. Given the length of time that the *CACNA1C* VNTR has existed, we expect that these large structural changes were rare and occurred early in the human lineages whose descendants are found today.

The complexity of the *CACNA1C* VNTR can be contextualized by comparison to other tandem repeats. Its 30-bp repeat unit size is close to the median (46^th^ percentile) for human-specific tandem repeat consensus units (81). The number of distinct *CACNA1C* VNTR Types was on par with the expected number of distinct alleles per VNTR genome-wide (7.5-16.7 alleles) (49). In contrast, its maximum length is ∼6 times longer than any of the 1,584 human-specific VNTR sequences identified in six long-read haplotype assemblies of Yoruban, Chinese, and Puerto Rican origin (81). Also, the average number of distinct repeat units per VNTR (8.97 ± 26.57 repeat units) was greatly exceeded by the 158 distinct repeat units of the *CACNA1C* VNTR (99^th^ percentile relative to other VNTRs). Comparisons to these genome-wide studies come with a caveat that their datasets were different subsets of the 155 haplotype assemblies described in this manuscript, capping their allelic diversity. Nonetheless, these analyses frame the *CACNA1C* VNTR within a genome-wide context as the human-specific VNTR with the greatest reported variability in length and the most diverse repeat unit structure.

The results described here address a tradeoff between conducting a single-locus analysis and consulting a pangenome (54) for complex genomic regions. While pangenome graphs can be referenced quickly, their accuracy is tied to the quality of the underlying alignment and the likely simplification of allelic complexity (41). In contrast, our detailed analysis of the *CACNA1C* VNTR establishes a template for teasing apart such complex GWAS associations. More work is needed to test the function of each candidate causal variant in disease cohorts and in a relevant cell type.

## MATERIALS AND METHODS

Additional information is available in ***SI Appendix***.

### Consensus *CACNA1C* VNTR sequences by Type

To align *CACNA1C* VNTR sequences in a unit boundary-aware manner, each VNTR sequence was encoded as a sequence of ASCII characters where each character represents one of 37 repeat units (**Fig. 1D**). To identify similar *CACNA1C* VNTR sequences, multiple sequence alignments were created using ‘`MAFFT –text –globalpair –maxiterate 1000`’. Final alignments for each Type were created using ‘`MAFFT --op 4 --text --globalpair`’.

A consensus sequence for each Type was defined from the most frequent unit at each alignment position. If the most frequent unit was an alignment gap with ≤ 65% frequency, the most frequent non-gap unit was used. In cases where two units were in equal frequency at a position, the more common unit among all 34,172 repeat units was chosen for the consensus.

### Calculating linkage disequilibrium between variable regions and nearby SNPs

We analyzed LD between variable regions and nearby SNPs in 35 individuals from HGSVC2 (52). Phased genotypes of VR1 and VR2 were included for the 34 HGSVC2 individuals contributing long-read assemblies to our dataset and one additional individual (HG02818) sequenced by both HGSVC2 and HPRC but whose VR alleles were identified using the HPRC assemblies. Custom VCF entries were created for VR1 and VR2 and inserted into the sorted file with phased variant calls. For each VR, the A allele (VR1A and VR2A) was encoded as the reference allele. VR1 was entered as a single biallelic variant and the four alleles of VR2 were split into three biallelic entries, one for each alternate allele (VR2B, VR2C, and VR2D). Positions in the reference genome for each entry were chosen within the reference *CACNA1C* VNTR region (chr12:2255791-2256090, GRCh38/hg38) on chromosome 12, starting at the first position (2255791–2255795). Only haplotypes with a Type 1 *CACNA1C* VNTR (n = 61) were encoded as either 0 or 1, indicating the reference or alternate VR allele respectively. Haplotypes with an unclassified VR or Type 2-7 sequence were indicated as missing (“.”).

SNP genotypes were taken from the HGSCV2 phased variant calls within the 1.4 Mb region surrounding the *CACNA1C* locus (chr12:1600001-3000000, GRCh38/hg38).

LD was calculated between each SNP-VR pair using ‘`vcftools --hap-r2`’.

## Data Availability

All data produced in the present study are available upon reasonable request to the authors

ftp://ftp.1000genomes.ebi.ac.uk/vol1/ftp/data_collections/HGSVC2/release/v1.0/assemblies

https://s3-us-west-2.amazonaws.com/human-pangenomics/index.html?prefix=working/

http://walters.psycm.cf.ac.uk/clozuk_pgc2.meta.sumstats.txt.gz

http://ftp.1000genomes.ebi.ac.uk/vol1/ftp/data_collections/HGSVC2/release/v2.0/integrated_callset/variants_freeze4_snv_snv_alt.vcf.gz

http://ftp.1000genomes.ebi.ac.uk/vol1/ftp/data_collections/1000G_2504_high_coverage/working/20201028_3202_phased/CCDG_14151_B01_GRM_WGS_2020-08-05_chr12.filtered.shapeit2-duohmm-phased.vcf.gz

https://www.internationalgenome.org/data-portal/sample

http://cdna.eva.mpg.de/neandertal/altai/AltaiNeandertal/bam/

http://ftp.eva.mpg.de/neandertal/Chagyrskaya/BAM/

http://cdna.eva.mpg.de/denisova/alignments/

http://ftp.eva.mpg.de/neandertal/Vindija/bam/Pruefer_etal_2017/

## ACKNOWLEDGEMENTS

This work was partially funded by National Institutes of Health (NIH) grants RM1HG009491, R35GM119703, and R01MH136353 (to M.T.M.) and R01NS125271 and R01MH071739 (to R.W.T.).

## SUPPLEMENTARY MATERIALS AND METHODS

### Data, Materials, and Software Availability

Download paths for long-read genome assemblies and whole-genome sequencing data are available in ***SI Appendix*** and **Dataset S1**. The R package for analyzing VNTRs in genome assemblies is available on GitHub at https://github.com/ramoya/analyzeRepeatSequences. GTEx v8 protected data is hosted in an AnVIL repository (https://gtexportal.org/home/protected-DataAccess). Access to GTEx protected data is available through the database of Genotypes and Phenotypes (dbGaP) (accession no. phs000424.v8). Summary statistics from the schizophrenia GWAS are available at http://walters.psycm.cf.ac.uk/clozuk_pgc2.meta.sumstats.txt.gz (6). Variant calls for HGSVC2 (http://ftp.1000genomes.ebi.ac.uk/vol1/ftp/data_collections/HGSVC2/release/v2.0/integrated_callset/variants_freeze4_snv_snv_alt.vcf.gz) (52) and 1000 Genomes individuals (http://ftp.1000genomes.ebi.ac.uk/vol1/ftp/data_collections/1000G_2504_high_coverage/working/20201028_3202_phased/CCDG_14151_B01_GRM_WGS_2020-08-05_chr12.filtered.shapeit2-duohmm-phased.vcf.gz) (86) are available through the 1000 Genomes FTP. WGS alignment files for archaic human individuals can be found at the following FTP sites: http://cdna.eva.mpg.de/neandertal/altai/AltaiNeandertal/bam/ (Denisova 5) (63), http://ftp.eva.mpg.de/neandertal/Chagyrskaya/BAM/ (Chagyrskaya 8) (61), http://cdna.eva.mpg.de/denisova/alignments/ (Denisova 3) (64), and http://ftp.eva.mpg.de/neandertal/Vindija/bam/Pruefer_etal_2017/ (Vindija 33.19) (62).

### Analysis of VNTR in long and short-read sequencing data

The CHM13 haploid genome assembly was downloaded from the Telomere-to-Telomere (T2T) GitHub site. Haplotype assemblies for 34 individuals were downloaded from the HGSVC2 (52) FTP site. In cases where two assemblies exist for an individual, assemblies using PacBio HiFi reads were preferred over PacBio continuous long reads (CLR). Haplotype assemblies for 43 individuals were downloaded from the HPRC (54) S3 bucket. 5 individuals were sequenced by both HGSVC2 and HPRC (HG00733, HG02818, HG03486, NA19240, NA24385). HGSVC2 assemblies were used for the individuals for which both projects had identical *CACNA1C* VNTR sequences (HG00733, HG03486, NA19240, NA24385). The HGSVC2 assembly of HG02818 had four contigs containing the consensus *CACNA1C* repeat unit, one of which matched the HPRC HG02818_paternal assembly. None of the other three contigs contained a full *CACNA1C* VNTR with unique flanking sequence, and thus the HPRC assembly was chosen for HG02818.

Corresponding short-read sequencing data (n = 70 individuals) were downloaded from the 1000 Genomes Project (86). Download paths are in **Dataset S1**. Assembly coordinates of each VNTR sequence are listed in **Dataset S3**.

Assemblies were scanned to identify contigs containing the previously identified *CACNA1C* VNTR consensus repeat unit (33). Tandem Repeats Finder (88) v4.09 was run on each resulting contig using parameters ‘`2 7 7 80 10 50 32 -m -f -d`’ to detect patterns near the size of the *CACNA1C* repeat unit. With a matching weight of 2 and a minimum alignment score of 50, at least 25 characters need to be aligned to the consensus repeat unit perfectly, which is permissive for a repeat unit of 30 bp. For this targeted approach to locate the *CACNA1C* VNTR, we used a maximum period size of 32 bp.

### Validation of haplotype assemblies over *CACNA1C* VNTR

We analyzed a subset of assemblies to confirm that PacBio reads support *CACNA1C* VNTR sequences found in the assemblies released by (52–54). Eight sequences were selected for analysis, one individual per Type (**Dataset S5**). For Type 1, the individual HG01071 was selected because it has two Type 1 *CACNA1C* VNTR sequences with heterozygous VR1B/VR1A and VR2B/VR2A. For Type 5, HG02818 was selected because it is the only individual with a Type 5 sequence on one allele and a non-Type 1 sequence (Type 2) on the other allele; also, its maternal haplotype has the longest *CACNA1C* VNTR sequence in this dataset. For Types 6 and 7, a single sequence was available. For Types 2, 3, and 4, a sequence was selected randomly.

A custom reference was created from each assembly’s *CACNA1C* VNTR sequence and 2.5 kb of flanking GRCh38/hg38 reference sequence. PacBio reads were downloaded from either the HGSCV2 FTP site or the HPRC S3 bucket (**Dataset S5**). For HPRC samples, whole-genome raw read BAM files were converted to FASTQ files. For each sample, PacBio reads were mapped to the relevant custom reference using minimap2 (89) and the preset value ‘`-x map-hifi`’. Unmapped reads were confirmed to not contain the *CACNA1C* VNTR consensus repeat unit and were dropped. Since the haplotype assignments for each PacBio read were not available for HPRC samples, we assigned reads to each haplotype based on either the pattern of mismatches over the VNTR or a heterozygous variant within the flanking sequence. In some cases we used a heterozygous variant within an expanded 5 kb flanking window to assign a read. Each BAM file was subset for the reads specific to one haplotype and a pileup was created using the Python module pysam for all positions of the custom reference.

### Assessing duplication within *CACNA1C* VNTR sequences

For each of the 26 gaps in the Type 1 consensus sequence, we extracted all non-gap sequences and summarized them into unique sequences (**Dataset S7**). We performed a census of divergence among these duplicated sequences and their sources (**Fig. S6**). Every gap in the consensus sequence was explained by a tandem duplication or a different equally optimal alignment. Five duplications were multi-allelic, yielding a total of 31 duplications observed in Type 1 sequences.

We also scanned each Type for larger duplications. An exemplar sequence with no duplications was defined for each VNTR Type. Using a sliding window of N repeat units, every overlapping window across the exemplar VNTR sequence was quantified in each sequence by counting exact matches using `grep`.

Because Types 1, 2 and 3 had similar repeat unit compositions (**Fig. 1G**), the Type 1 consensus sequence was defined as an exemplar sequence for all three. The width of the sliding window for Types 1, 2, and 3 was 6 repeat units.

For Types 4, 6, and 7, their consensus sequences were defined as exemplar sequences and scanned for duplications within themselves. The width of the sliding window for Types 4, 6, and 7 was 6 repeat units.

Because the Type 5 consensus sequence contained duplications itself, the Type 5 exemplar sequence was derived from the HG00735_paternal sequence by excluding 5 small insertions and 3 single repeat unit insertions present as gaps in the multiple sequence alignment of all four Type 5 sequences (**Fig. 1E**). The width of the sliding window for Type 5 was 8 repeat units.

Type 5 was also distinguished from Types 1-3 by the high density of repeat unit mismatches throughout the sequence relative to the exemplar sequence. Relative to HG00735_paternal, the other three Type 5 sequences had repeat unit mismatches that occur across the entire sequence.

In contrast, Types 1-3 had repeat unit mismatches concentrated in a 690-bp segment overlapping VR2, leading us to reason that this segment had a different repeat unit composition in Types 1, 2, and 3. Thus we analyzed the VR alleles of Type 2 and 3 sequences. Consistent with their derivation from Type 1 and with each sequence’s duplication dosage, Type 2 and 3 had multiple copies of known VR alleles (**Fig. S13**, **Supplementary Materials and Methods**).

### Identifying variable repeat regions and alleles

Variability at each position in the multiple sequence alignment was computed as Shannon’s uncertainty 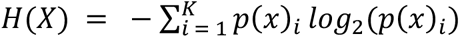 where *p_i_* is the fraction of repeat units of unit type *i* and *K* is the number of different repeat units at position *X*. Gaps are included in the calculation. *H*(*X*) was normalized as 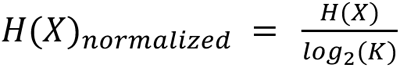 and a smoothing filter was applied using the R package ksmooth. Variable regions were defined where 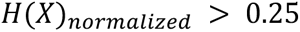 and length > 7 aligned repeat units. Variable region sequences were inspected manually and narrowed by a maximum of 2 repeat units on either end (***SI Appendix***, **Fig. S7**).

For each variable region, aligned sequences were extracted from Type 1 sequences and partitioned into alleles. First, a consensus sequence for one allele was defined as the most common VR sequence (***SI Appendix***, **Dataset S8**, **Dataset S9**). Hamming edit distances in repeat units were calculated between the consensus sequence and each unique VR sequence using `StrDist` in the R package DescTools. A consensus sequence for the second allele was defined as the next most common VR sequence with a large edit distance from the first allele. Similarly, edit distances (in repeat units) were calculated between the second allele and each unique VR sequence. Partitioning VR sequences into alleles was done using edit distance thresholds. For example, a threshold *t* was chosen such that a sequence belonging to the A allele had less than *t* mismatches (in repeat units, including gaps) to the consensus A sequence and greater than *t* mismatches to the consensus B sequence. For VR1 t = 7.5, for VR2A t = 10, and for VR2B t = 12.5. If VR sequences clustered into more than two groups, the other groups were manually defined as rarer VR alleles. Unclassified VR sequences fell outside the selected edit distance thresholds.

For nucleotide-level alignments (***SI Appendix***, **Fig. S8B**, **Fig. S9B**), each aligned VNTR sequence was converted from encoded characters to nucleotides while preserving the positions of alignment gaps. Global alignments were adjusted using `pairwiseAlignment` from the R package Biostrings to account for re-incorporation of non-30-bp repeat units.

To identify VR alleles in Type 2 and Type 3 sequences, *CACNA1C* VNTR sequences were scanned for matches to each consensus VR allele allowing for maximum 2 mismatches (in repeat units) using `vmatchPattern` from the R package Biostrings.

### Analysis of VRs in Type 2 and Type 3 sequences

We identified only two VR1 sequences and one VR2 sequence within Types 2 and 3 that had been previously identified in Type 1 (**Fig. S13A** and **B**). However, all except one VR2 sequence corresponded to known VR1 and VR2 alleles. Types 2 and 3 harbored two or three complete duplications of VR1, and VR2 was partially duplicated in Type 2 (**Fig. 2B**, **Fig. S13A** and **B**). Interestingly, VR1 shared a right breakpoint with Duplication 2 and the Triplication in Type 3. VR1 in Type 1 sequences is a duplication of its right flanking sequence with some divergence at the same breakpoint (**Fig. 2B**, Duplication 5), suggesting this site might be prone to instability. When VR1A was duplicated, one copy typically matched the consensus while the other had more mismatches. VR1B sequences in Type 2 did not show this pattern; instead they were exact copies, perhaps because VR1B is shorter and therefore has less scope to accumulate mutations. VR1 and VR2 alleles had the same correlation in Type 2 and 3 as in Type 1. VR1A was found in the same sequences as VR2C, while VR1B was found in the same sequences as VR2B. Type 2 represented two of the four possible versions of Type 1 sequences, while Type 3 only represented one version (**Fig. S13C** and **D**). Thus, Type 2 and Type 3 are the result of large duplications within a subset of Type 1 sequences.

### GWAS data retrieval

Summary statistics of the CLOZUK+PGC2 schizophrenia meta-analysis and the subset of highquality imputed SNPs (6) were obtained from the Psychiatric Genomics Consortium repository. SNP coordinates were converted from GRCh37/hg19 by looking up GRCh38/hg38 coordinates in dbSNP using the rsID. Odds ratio (OR) was used to identify risk (OR > 1) and protective (OR < 1) alleles. Fine-mapped SNPs were obtained from Supplementary Table 11 of the same study (6) and similarly converted to GRCh38/hg38.

### Analysis of GTEx eQTLs

eQTLs from the v8 analysis freeze were downloaded from the GTEx Portal on 09/20/2017. Analysis was limited to significant associations with ENSG00000151067.21 (Ensembl) within chr12:1600001-3000000 (GRCh38/hg38).

For eQTLs colocalized with schizophrenia associations, the direction of eQTL effect was defined as the risk allele relative to the protective allele. The slope of the linear regression was inverted for associations where the reference allele matched the risk allele, which negated the default direction of the alternate allele relative to the reference allele.

To estimate the magnitude of *CACNA1C* expression change associated with SNPs in intron 3, software for calculating the log allelic fold change (aFC) was downloaded from the aFC GitHub site. aFC is equivalent to the expected log-fold expression ratio of the individuals homozygous for the alternate allele to those homozygous for the reference allele of an eQTL (57). aFC was calculated using ‘`aFC.py --min_samps 2 --min_alleles 1 --log_xform 1 --log_base 2`’ for eQTLs in cerebellar hemisphere. Normalized expression values, covariates, and phased variant calls were included. Phased variant calls for 838 GTEx individuals were obtained from dbGaP (release v8, GTEx_Analysis_2017-06-05_v8) on 01/12/2019.

### Calculating *CACNA1C* VNTR length by WGS coverage depth

Local sequencing coverage depth was used to estimate *CACNA1C* VNTR length. Average depth over three regions (left flanking 10 kb: chr12:2245791-2255790, right flanking 10 kb: chr12:2256091-2266090, *CACNA1C* VNTR region: chr12:2255791-2256090, GRCh38/hg38) was calculated using ‘`samtools depth -a`’. *CACNA1C* VNTR length was estimated by computing the ratio of coverage depth over the VNTR to the coverage depth over each flanking sequence, then taking the average of these two ratios (**Fig. S16A**). This value was scaled by a conversion factor 300/30 to account for the length of the collapsed repeat in the GRCh38/hg38 reference sequence. The output length estimate corresponds to the average of both alleles (in number of repeat units) within a diploid genome.

BAM files containing chromosome 12 for four archaic human individuals were downloaded. *CACNA1C* VNTR lengths were estimated as before, using UCSC liftOver to identify the corresponding regions in GRCh37/hg19.

The coverage depth distribution limited to 20-bp mappable regions is similar to the coverage depth over the whole Denisova 3 genome (64), indicating that repetitive regions may be sequenced at a similar rate as non-repetitive sequences in ancient hominin genomes although no global analysis can show this directly. Coverage depth for Denisova 3 is slightly higher at low G+C regions relative to modern human genomes (64). But the *CACNA1C* VNTR consensus repeat unit is 47% GC (n = 14/30 nucleotides), and the left and right flank are 49% and 44%, respectively, so this is not expected to change the coverage ratios significantly relative to modern human genomes.

## SUPPLEMENTARY FIGURES

**Fig. S1.**
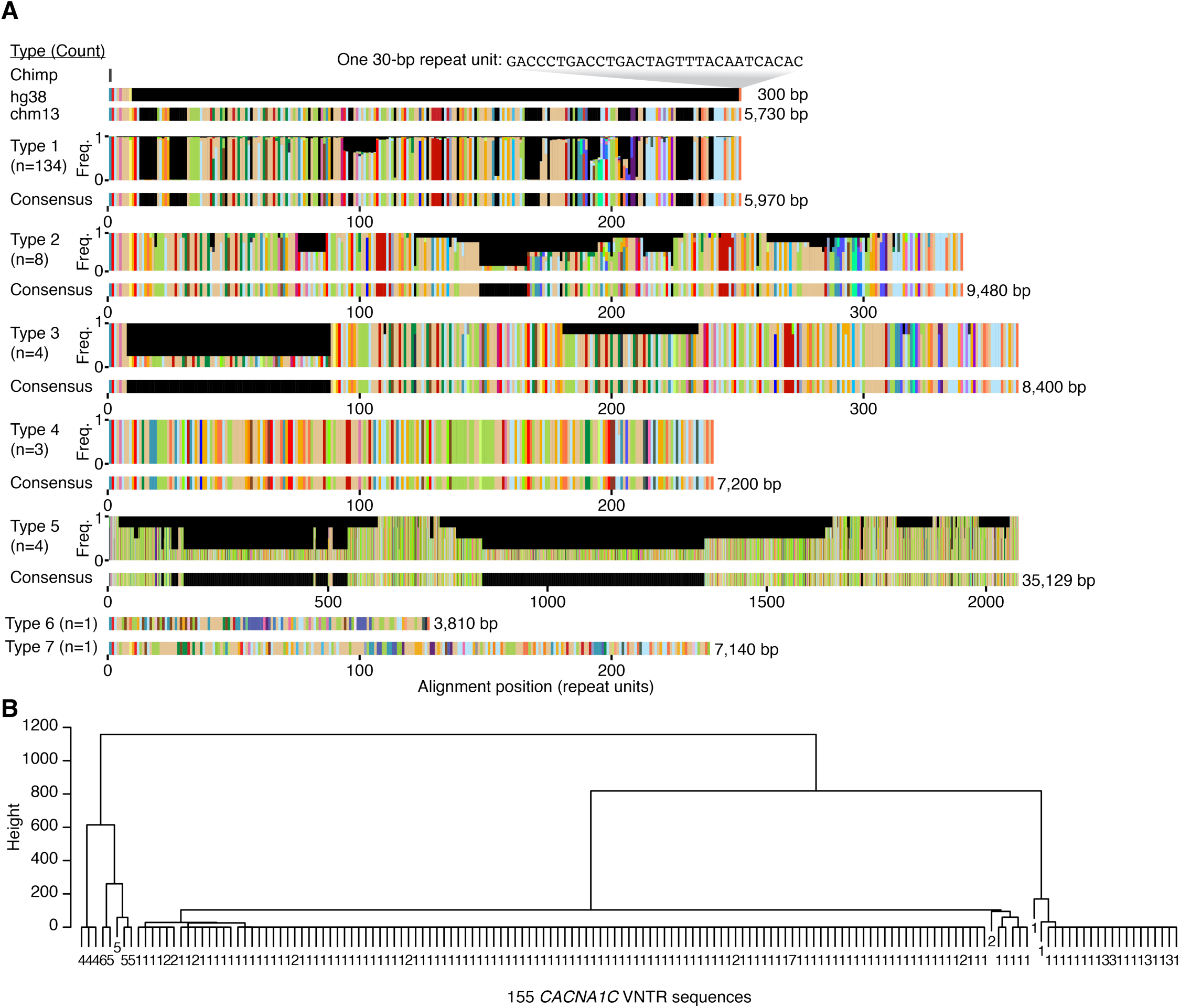
Definition of 7 *CACNA1C* VNTR Types. (***A***) Chimp (dark gray), human genome (hg38), and T2T (chm13) reference sequences are shown at top. For each VNTR Type, repeat unit frequencies (y-axis) per position (x-axis) are shown above a consensus sequence. Type 5 has a different x-axis scale than the other Types. Numbers in parentheses indicate VNTR Type frequency among 155 sequences. Unaligned lengths (bp) are shown to the right of each consensus. Type 6 and Type 7 each represent single VNTR sequences dissimilar to any other Type. (***B***) Dendrogram representing Ward’s minimum variance agglomerative clustering of the multiple sequence alignment of 155 VNTR sequences. Each leaf is one repeat sequence labeled by its VNTR Type.

**Fig. S2.**
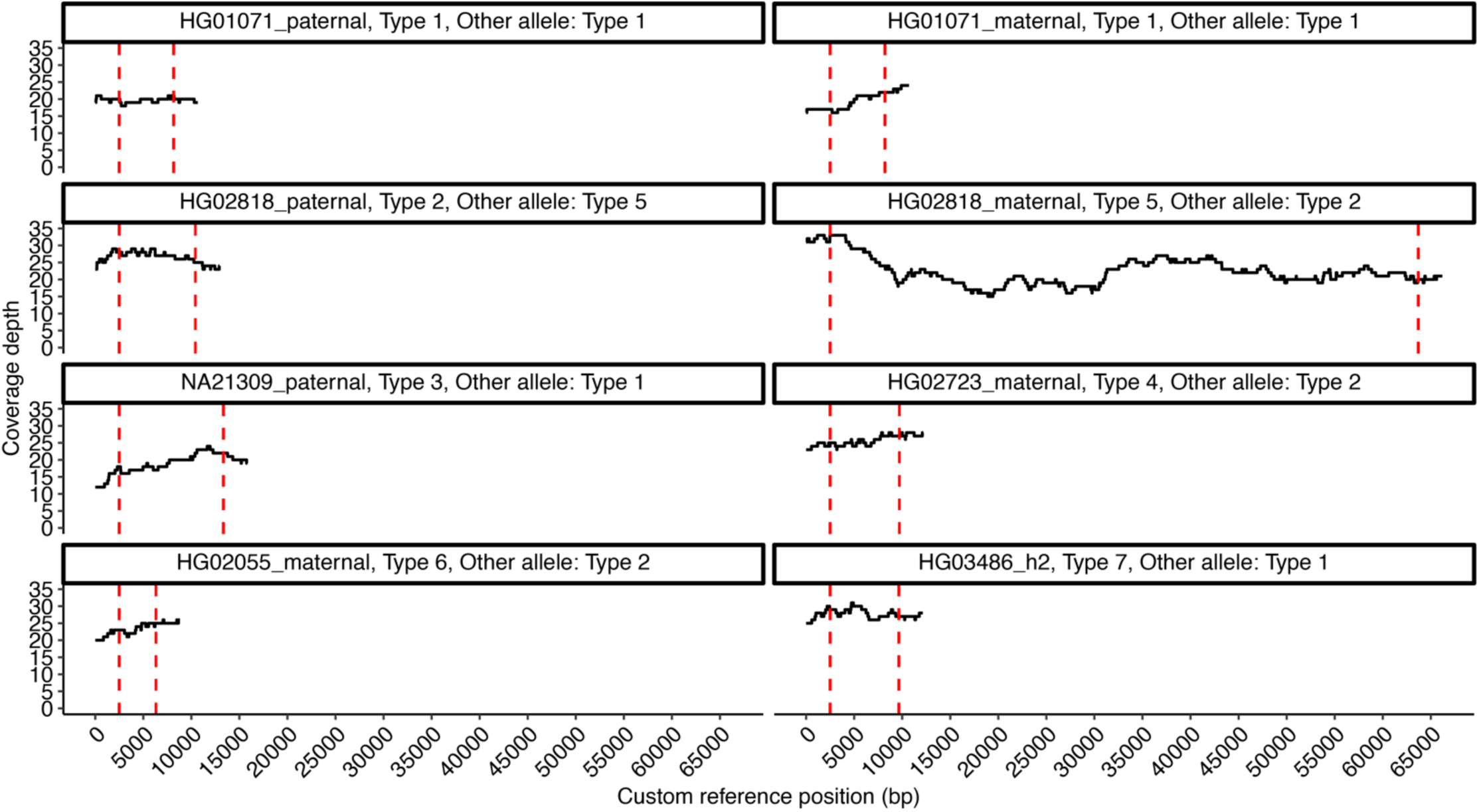
Coverage depth across *CACNA1C* VNTR assemblies. Coverage depth over eight *CACNA1C* VNTR sequences encompassing each Type and 2.5 kb of flanking GRCh38/hg38 reference sequence. The position of the VNTR is between two vertical dashed red lines. Details of the sequences analyzed are in **Dataset S5**.

**Fig. S3.**
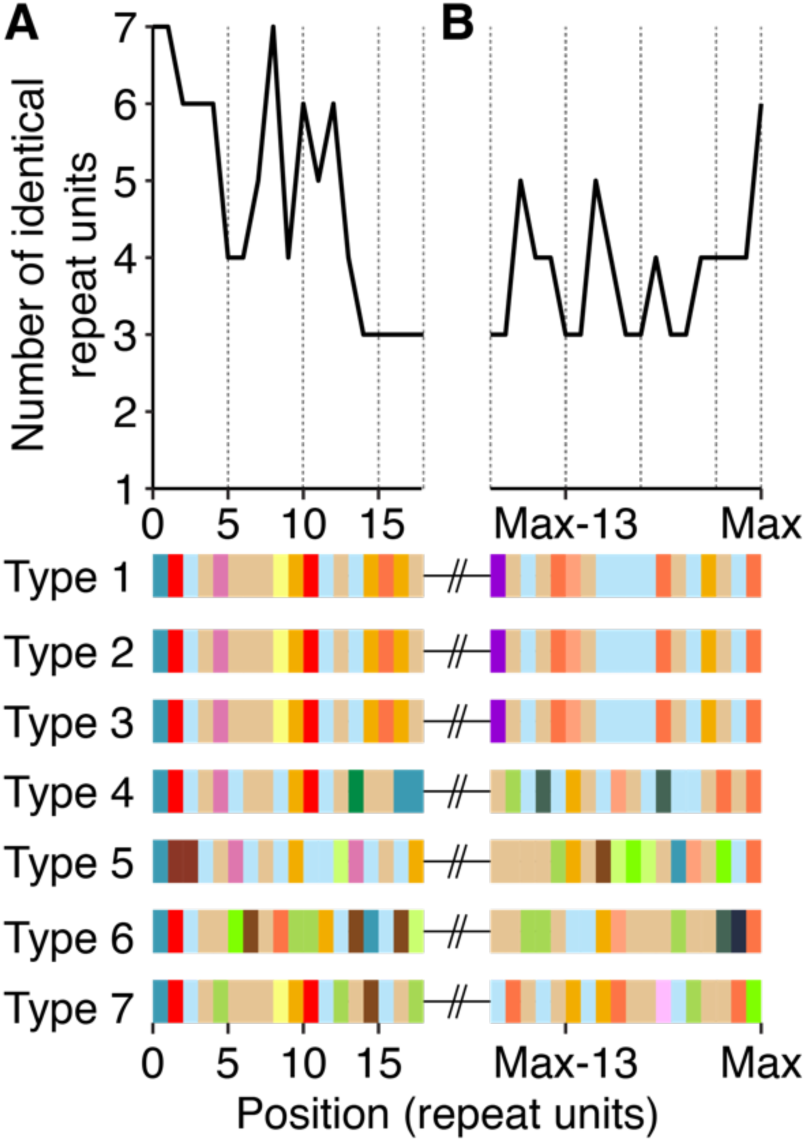
Comparison of starting and ending repeat units across Types. (***A*** and ***B***) Count of identical repeat units across the first (***A***) and last (***B***) 18 repeat units of each Type consensus sequence. Colors are as in Fig. 1D.

**Fig. S4.**
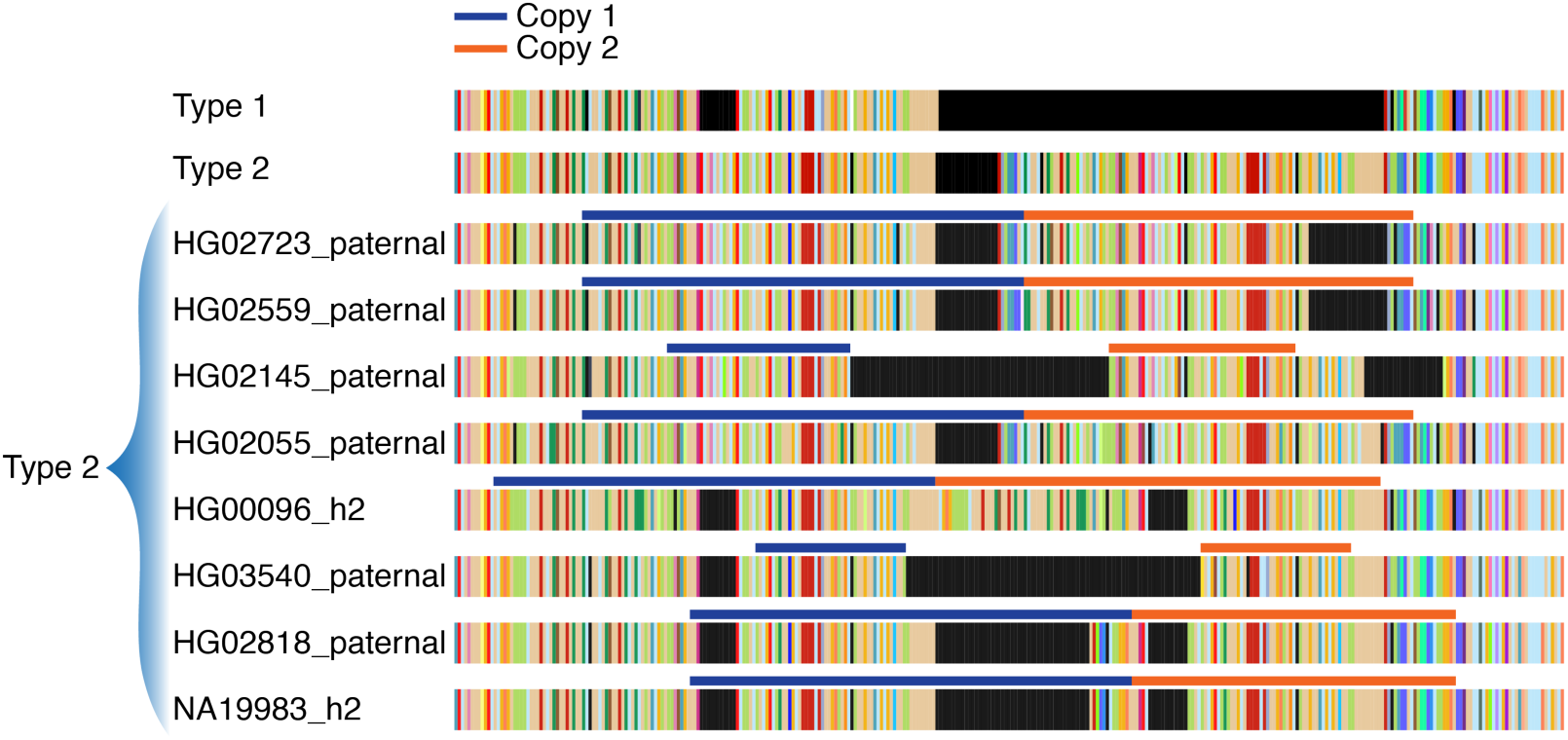
Duplications in Type 2 sequences. Breakpoints of the duplication within each Type 2 sequence. Type 1 and Type 2 consensus sequences are shown above.

**Fig. S5.**
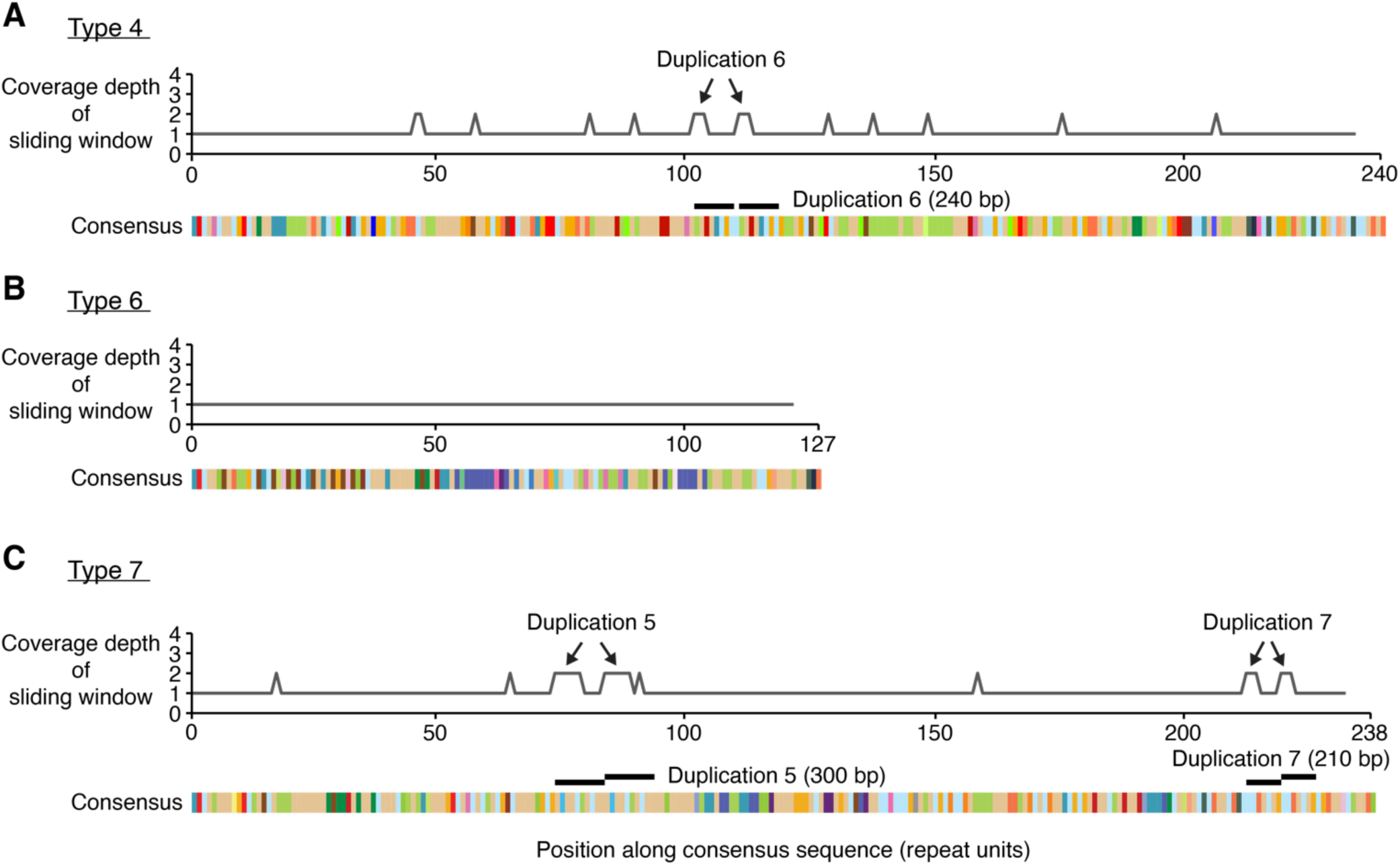
Duplication scans for Types 4, 6, and 7. Coverage depth (y-axis) of a sliding window (width = 6 repeat units) across the consensus sequences of Type 4 (***A***), Type 6 (***B***), and Type 7 (***C***). Pairs of equally-sized peaks identify three tandem duplications, numbered by decreasing size: Duplication 6 in Type 4, and Duplications 5 and 7 in Type 7. Duplication 6 is not exactly in tandem; it is separated by one repeat unit. Other smaller regions in Type 4 and Type 7 consensus sequences have a coverage depth of 2 at multiple locations, but are not contiguous and therefore do not reflect tandem duplications. Type 6 has coverage depth of 1 of the sliding window across its whole sequence.

**Fig. S6.**
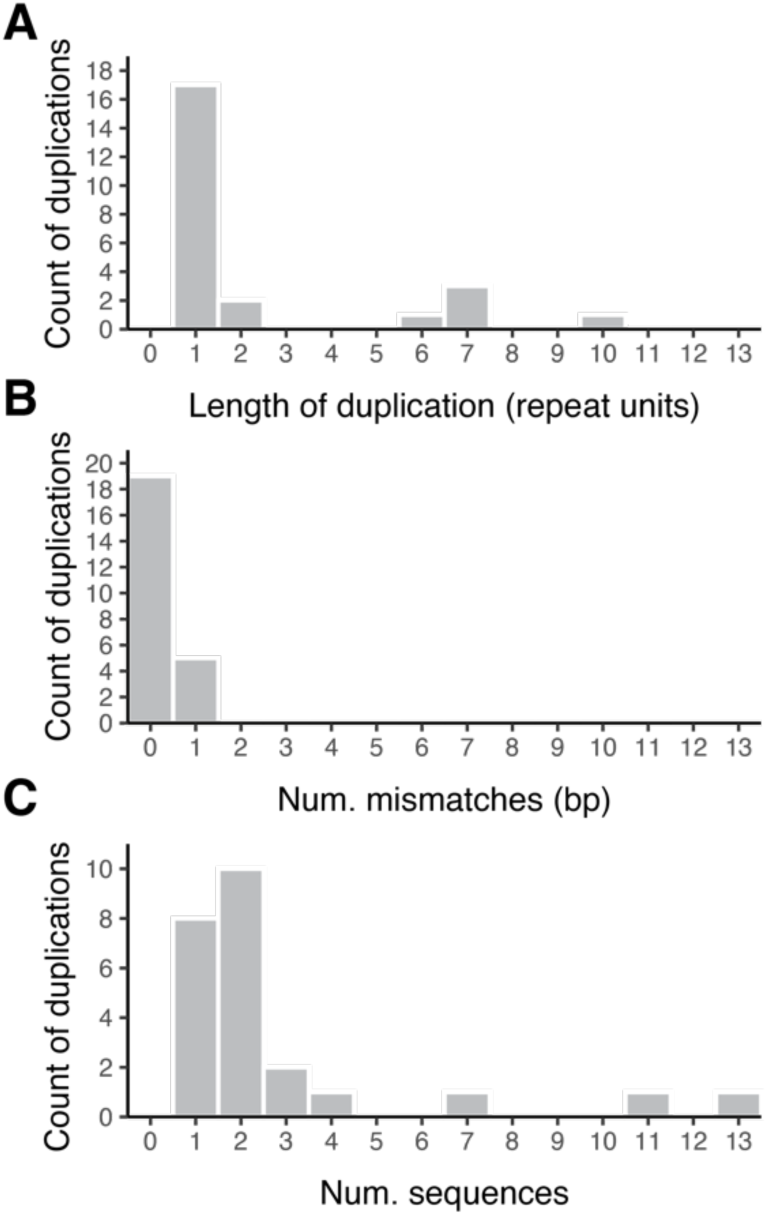
Duplications in Type 1 sequences. (***A***) Length of each duplication (n = 31) in repeat units. (***B***) Number of mismatches (in nucleotides) between each duplication’s source and destination sequence. (***C***) Number of Type 1 sequences that each duplication occurred in.

**Fig. S7.**
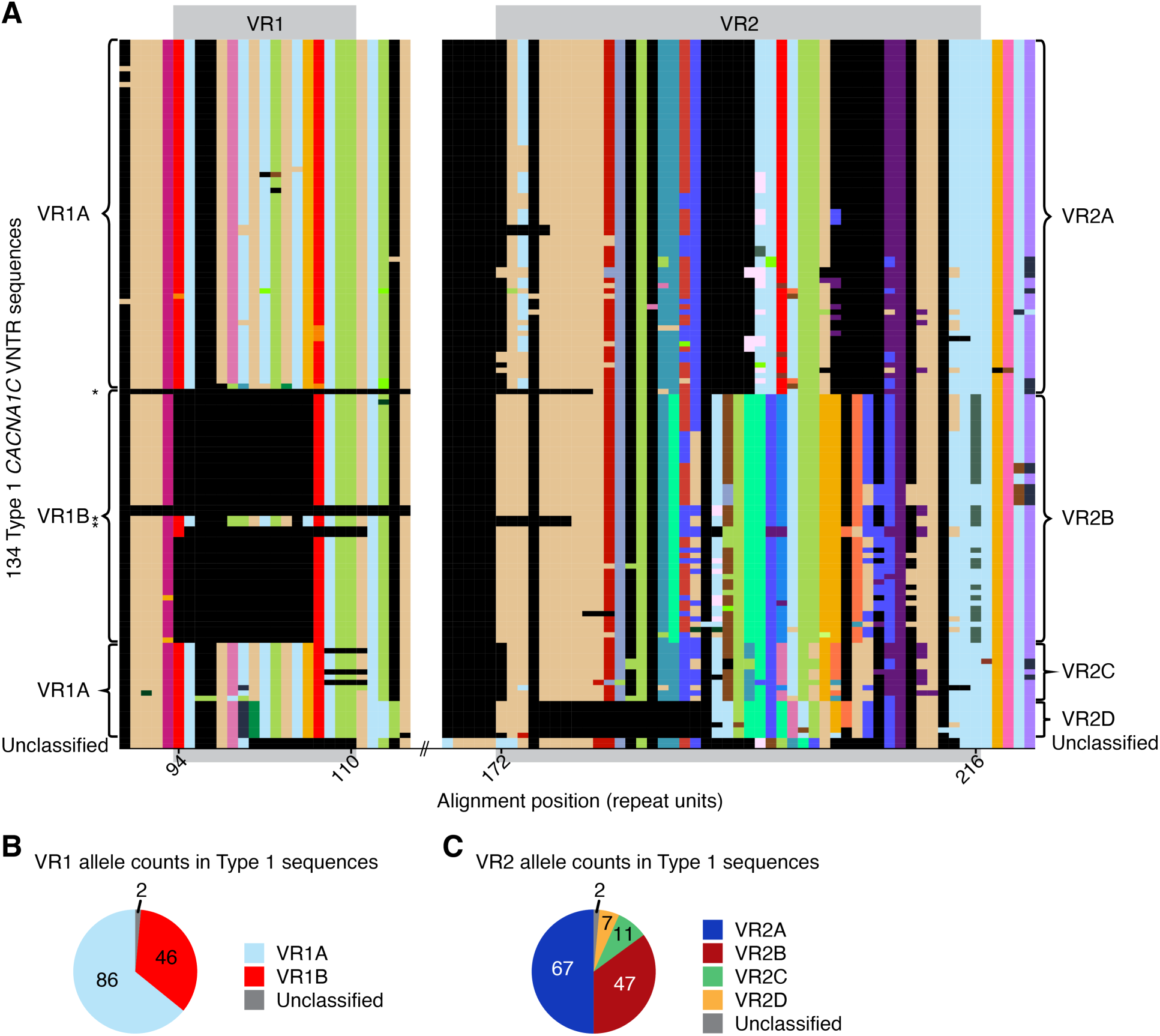
Expanded view of variable regions. (***A***) Enlargement of VR1 and VR2 in the alignment of Type 1 sequences (Fig. 3B). Constant regions of 5 repeat units flank each VR. Sequences are ordered by VR2 allele classification (right), revealing near-complete correspondence with VR1 allele classification (left); black asterisks (left column) denote the only three VR1 sequences that deviate from this pattern. (***B***) VR1 allele counts in 134 Type 1 sequences. (***C***) VR2 allele counts in 134 Type 1 sequences.

**Fig. S8.**
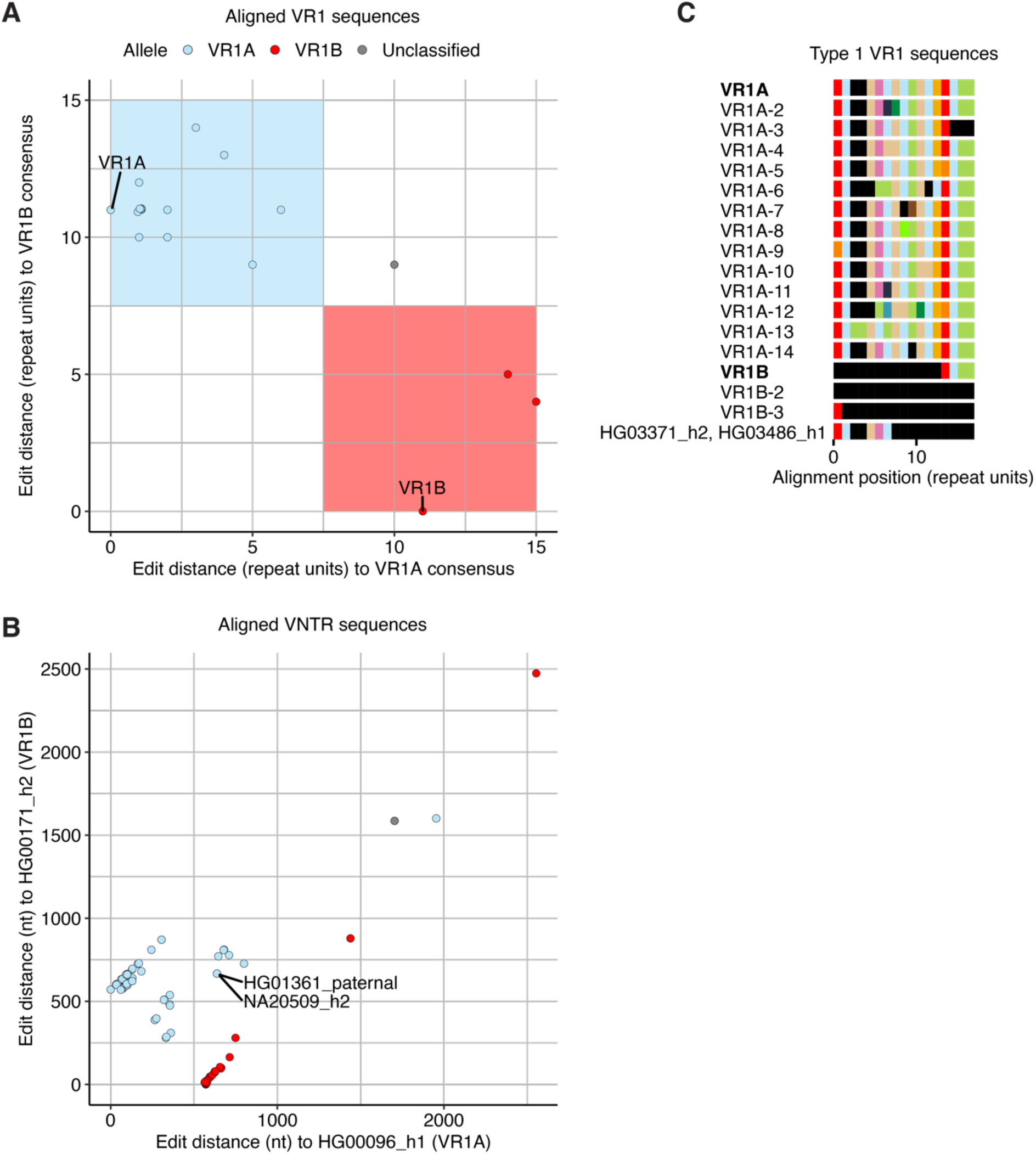
VR1 allele definition. (***A***) Scatterplot of edit distances (in repeat units) to the VR1B consensus vs. the VR1A consensus. Each point represents one of the 18 distinct VR1 sequences. Overlapping points are plotted with a small amount of jitter. Shaded areas indicate thresholds used to partition sequences into alleles. (***B***) Scatterplot of edit distances (at the nucleotide level) to two exemplar full *CACNA1C* VNTR sequences. Each point represents a full, aligned Type 1 sequence (n = 134) and its coordinates are edit distances to a VNTR sequence with the consensus VR1B allele (HG00171_h2, y-axis) and a VNTR sequence with the consensus VR1A allele (HG00096_h1, x-axis). VR1 alone does not explain the clustering of these sequences. However, two exceptional *CACNA1C* VNTR sequences with VR1A and VR2B (HG01361_paternal, NA20509_h2) cluster these sequences by their VR1 allele (see **Fig. S7**, asterisks). (***C***) Consensus VR1 sequences and their forms (see **Dataset S8** for details) identified in Type 1 sequences. Consensus alleles are in bold. Non-consensus VR1 sequences classified to each allele are numbered and listed below the consensus. Three *CACNA1C* VNTR sequences (HG00732_h1, HG00733_h2, and NA19650_h1) contained large deletions that included VR1, resulting in the VR1 sequence VR1B-2. The unclassified VR1 sequence is named by the assembly IDs it is identified in.

**Fig. S9.**
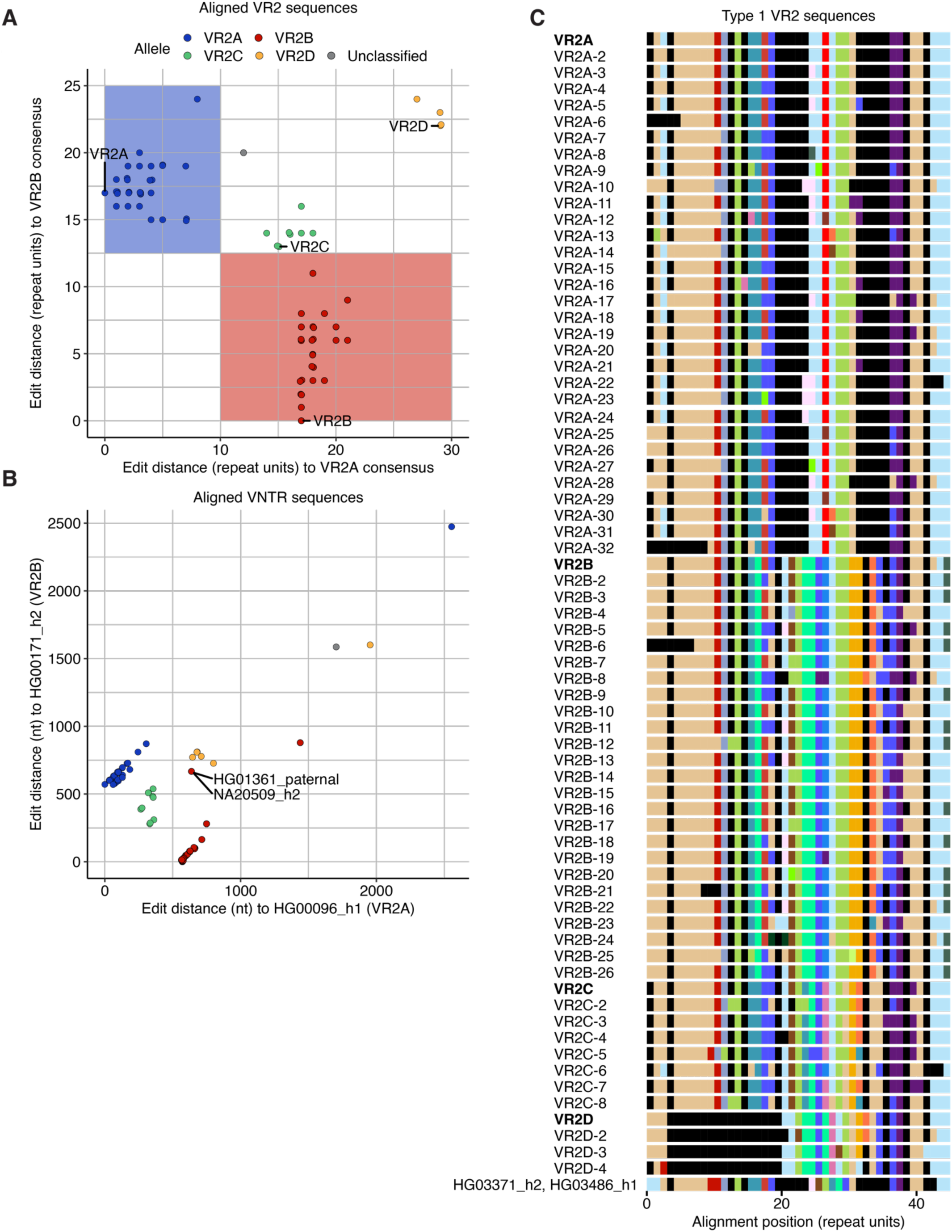
VR2 allele definition. (***A***) Scatterplot of edit distances (in repeat units) to the VR2B consensus vs. the VR2A consensus. Each point represents one of the 71 distinct VR2 sequences. Overlapping points are plotted with a small amount of jitter. Shaded areas indicate thresholds used to partition sequences into two common alleles. The two rarer alleles (VR2C and VR2D) were defined manually. (***B***) Scatterplot of edit distances (at the nucleotide level) to two exemplar full *CACNA1C* VNTR sequences. Each point represents a full, aligned Type 1 sequence (n = 134) and its coordinates are edit distances to a VNTR sequence with the consensus VR1B allele (HG00171_h2, y-axis) and a VNTR sequence with the consensus VR1A allele (HG00096_h1, x-axis). Sequences cluster according to VR2 allele. However, two *CACNA1C* VNTR sequences (HG01361_paternal, NA20509_h2) with a VR2B allele (VR2B-6) inappropriately cluster near sequences with VR2D alleles because they have VR1A (VR1A-6); most sequences with VR2B have VR1B. (***C***) Consensus VR2 sequences and their forms (see **Dataset S9** for details) identified in Type 1 sequences. Consensus alleles are in bold. Non-consensus VR2 sequences classified to each allele are numbered and listed below the consensus. One *CACNA1C* VNTR sequence (NA19650_h1) has a large deletion that includes VR1 and part of VR2, resulting in the VR2 sequence VR2A-32. The unclassified VR2 sequence is named by the assembly IDs it is identified in.

**Fig. S10.**
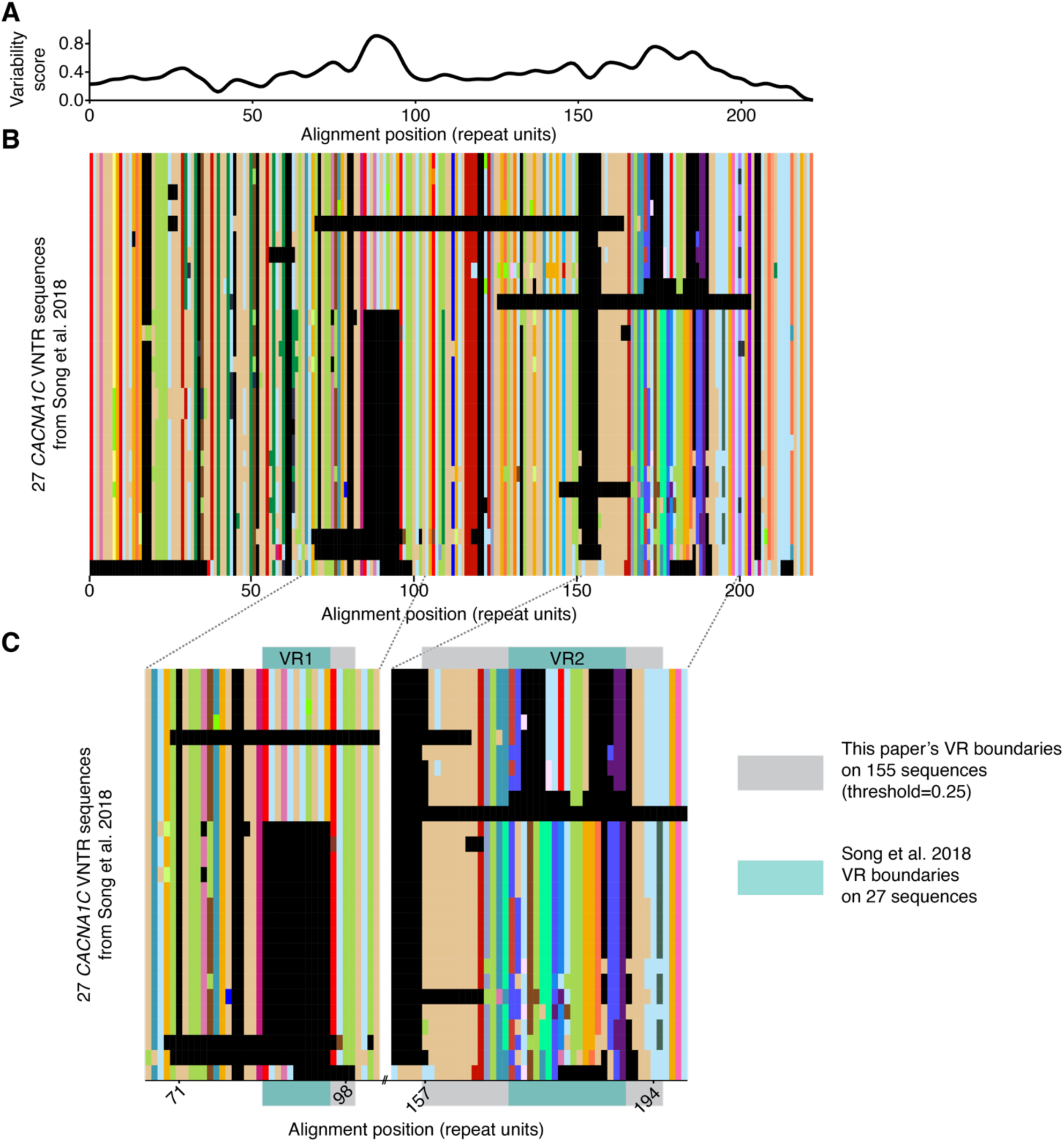
Comparison to previously-identified VR boundaries. (***A***) Repeat unit variability, shown as normalized Shannon’s uncertainty H(x), above the (***B***) multiple sequence alignment of 27 *CACNA1C* VNTR sequences (GenBank accessions MH645925-MH645951) that were long-read sequenced following PCR amplification and size-selection (33). Variable regions are defined at H(x) > 0.25 and length > 7 aligned repeat units. (***C***) Expanded view of variable regions comparing their previously described boundaries (33) to the boundaries defined in this paper.

**Fig. S11.**
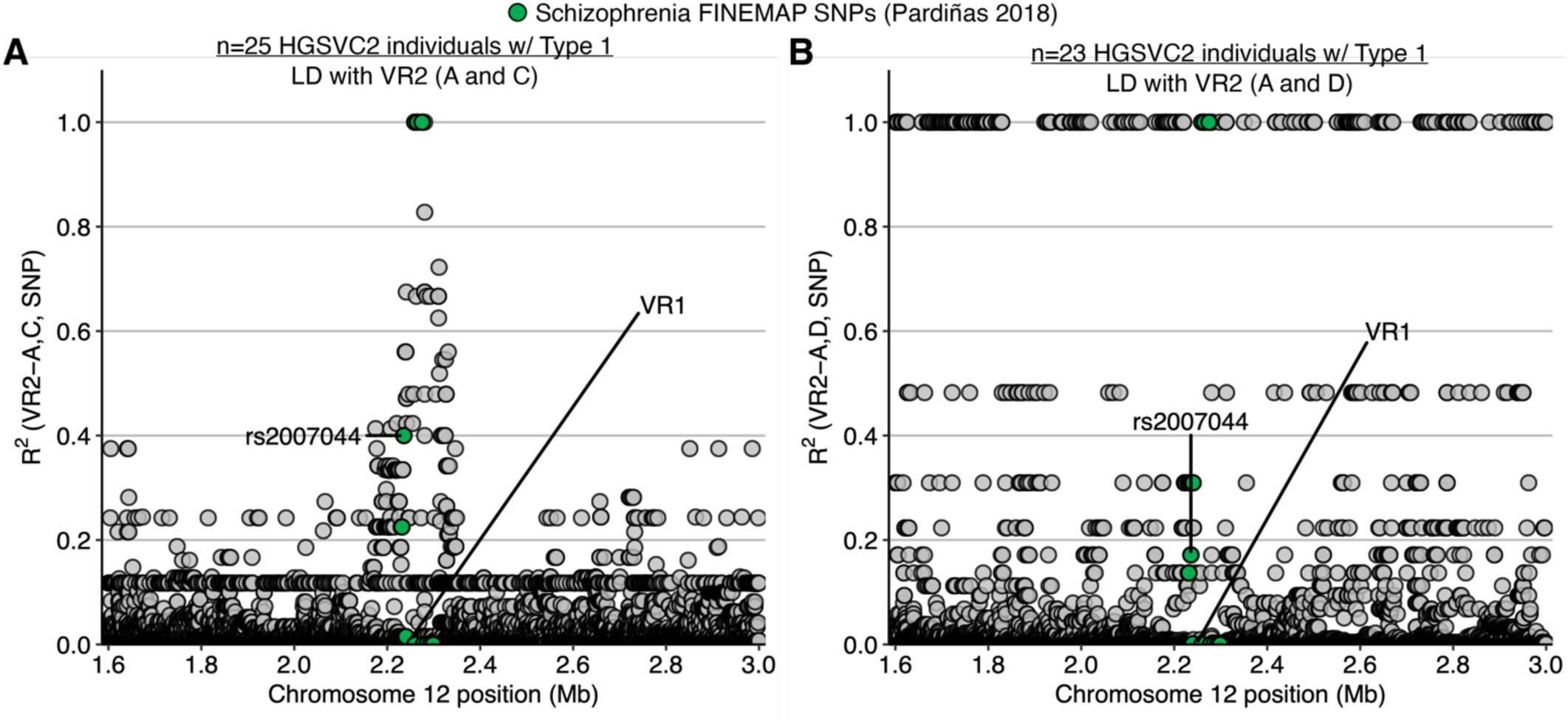
Linkage disequilibrium between schizophrenia association and rarer VR2 alleles. (***A*** and ***B***) Linkage disequilibrium (LD, y-axis) between surrounding SNPs (x-axis) and VR2C (***A***) and VR2D (***B***). Fine-mapped schizophrenia SNPs are highlighted in green. VR2C (frequency = 0.08) shows a similar LD pattern as VR2B with fine-mapped schizophrenia SNPs. Though observed infrequently (frequency = 0.05), VR2D displays a different LD structure with nearby SNPs.

**Fig. S12.**
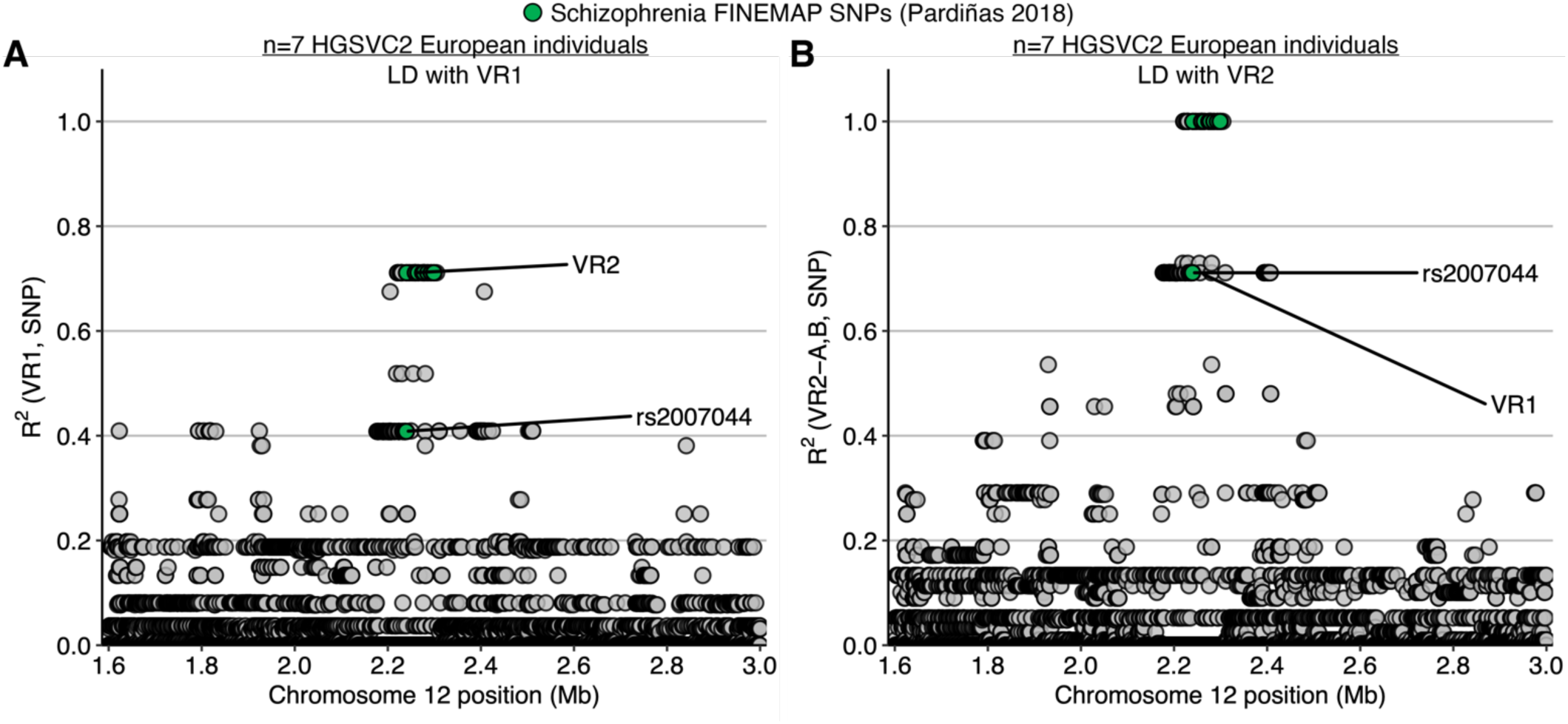
Linkage disequilibrium between VR2 and schizophrenia SNPs in Europeans. (***A*** and ***B***). Linkage disequilibrium (LD, y-axis) between VR1 (***A***) or VR2 (***B***) and surrounding SNPs in 7 HGSVC2 European individuals. Fine-mapped schizophrenia SNPs are highlighted in green. LD was calculated for VR2A and VR2B alleles in (***B***).

**Fig. S13.**
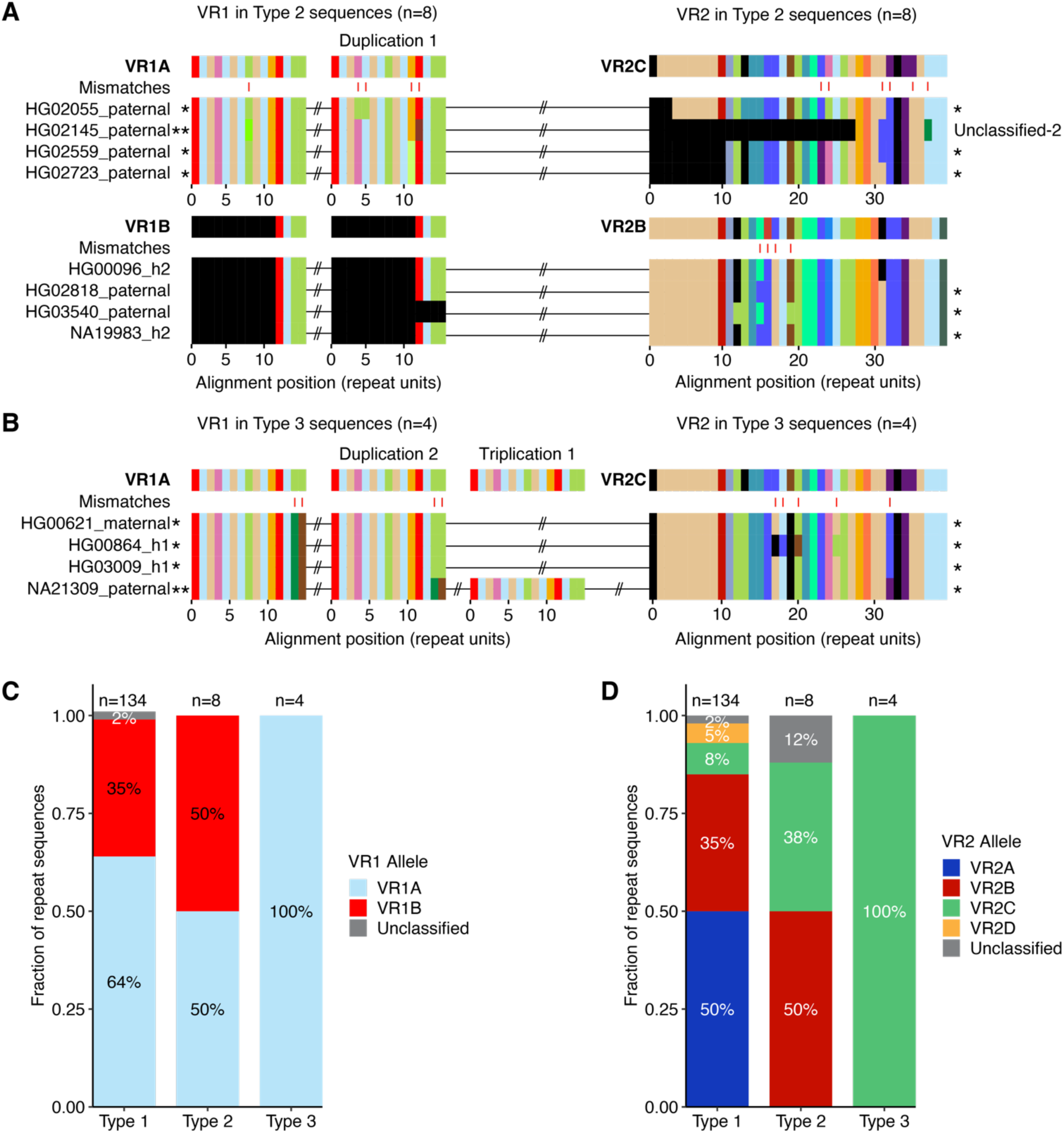
Variable region alleles in Type 2 and Type 3 *CACNA1C* VNTR sequences. (***A*** and ***B***) VR sequences in Type 2 (***A***) and Type 3 (***B***) sequences. Each VR is aligned to its consensus. In Type 2, VR1 is duplicated and is found as VR1A and VR1B in equal proportion; VR2 is found as VR2B and VR2C in nearly equal proportion except for one unclassified sequence. In Type 3, VR1 exists as VR1A only, and VR2 exists as VR2C. Asterisks indicate sequences unique to Type 2 and 3. The number of asterisks is proportional to the copy number VR sequences unique to Type 2 and 3. Positions with a mismatched repeat unit in any sequence relative to the consensus are shown with red ticks; mismatches between a gap and a repeat unit are not shown. (***C*** and ***D***) Frequency of VR1 (***C***) and VR2 (***D***) alleles in Type 1, 2, and 3 sequences. Only the primary copy is counted for sequences with a duplicated or triplicated VR.

**Fig. S14.**
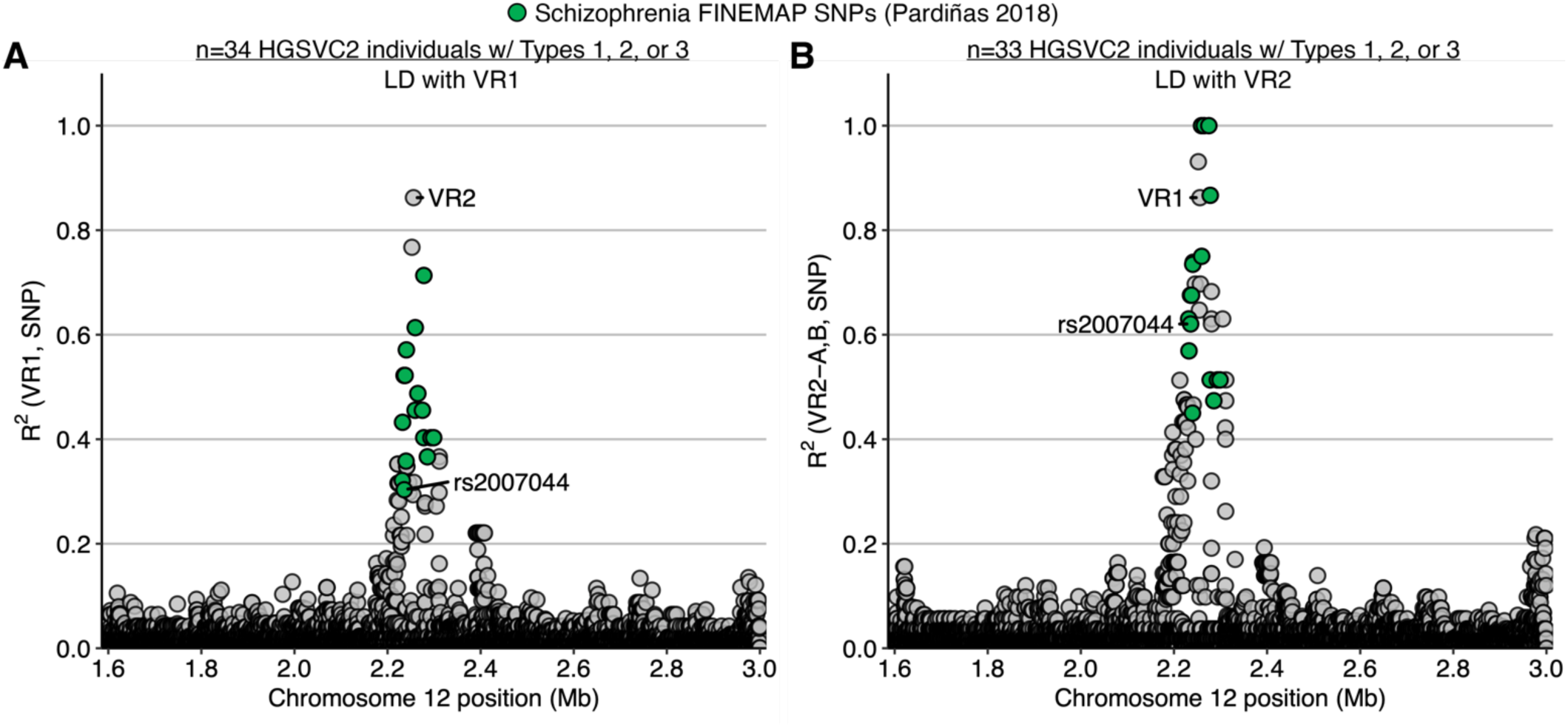
Linkage disequilibrium between schizophrenia association and Type 1-3 VRs. (***A*** and ***B***) Linkage disequilibrium (LD, y-axis) between surrounding SNPs (x-axis) and VR1 (***A***) and VR2 (***B***) in HGSVC2 individuals with at least one Type 1, 2, or 3 sequence. Fine-mapped schizophrenia SNPs are highlighted in green. LD was calculated for VR2A and VR2B alleles in (***B***).

**Fig. S15.**
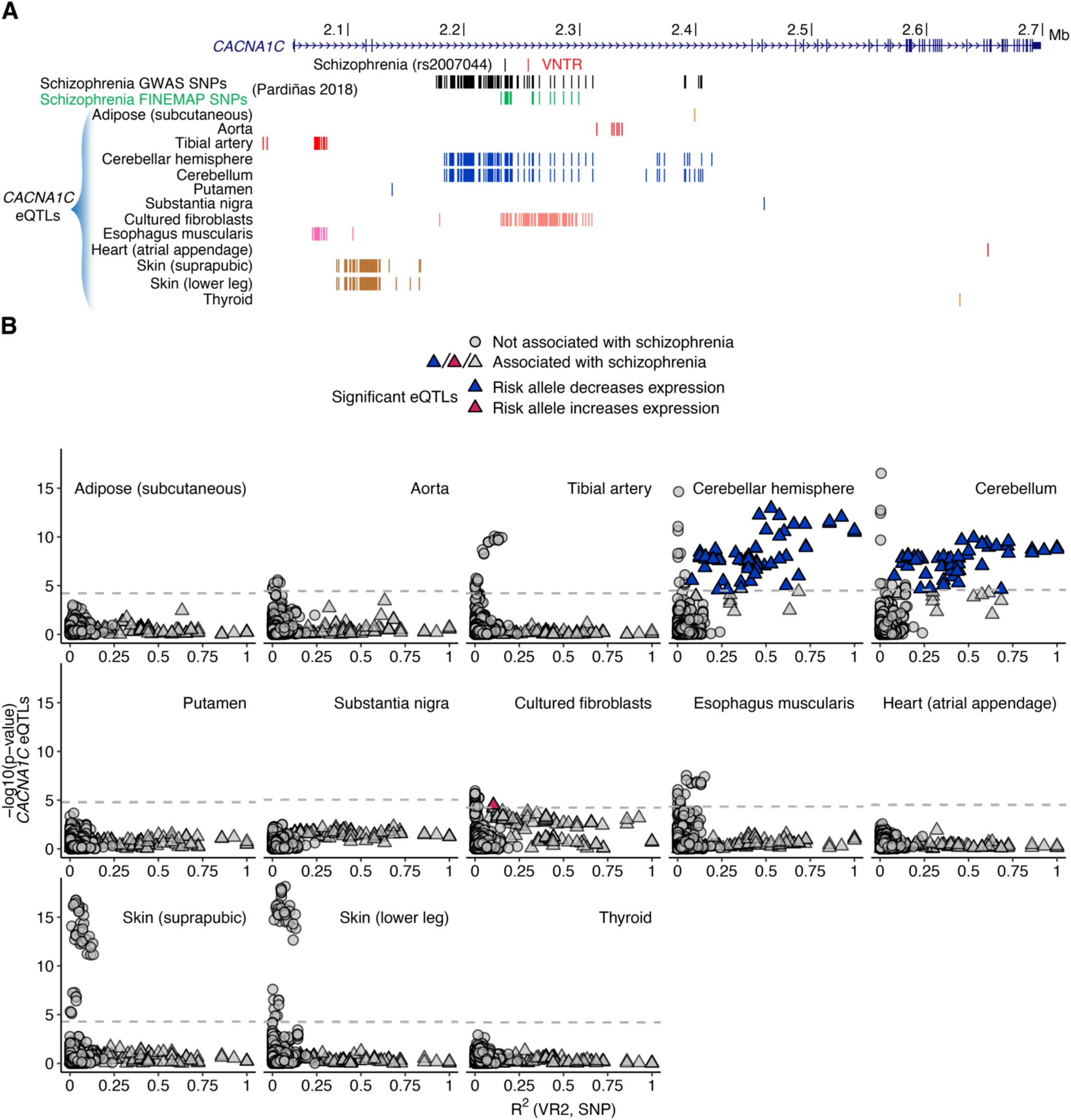
VR2 linkage disequilibrium with GWAS and eQTL SNPs per tissue. (***A***) *CACNA1C* eQTLs by GTEx tissue. (***B***) Analysis of the relationship between VR2 and eQTLs in GTEx tissues with at least one *CACNA1C* eQTL. Only cerebellar hemisphere and cerebellum had an eQTL signal associated with schizophrenia and VR2 (blue). Dashed gray line indicates eQTL *P* value threshold. Significant *CACNA1C* eQTLs for adipose (subcutaneous) (n = 1), putamen (n = 1), substantia nigra (n = 1), heart (atrial appendage) (n = 1), and thyroid (n = 2) are eliminated after intersecting data with schizophrenia GWAS and LD results and thus not shown.

**Fig. S16.**
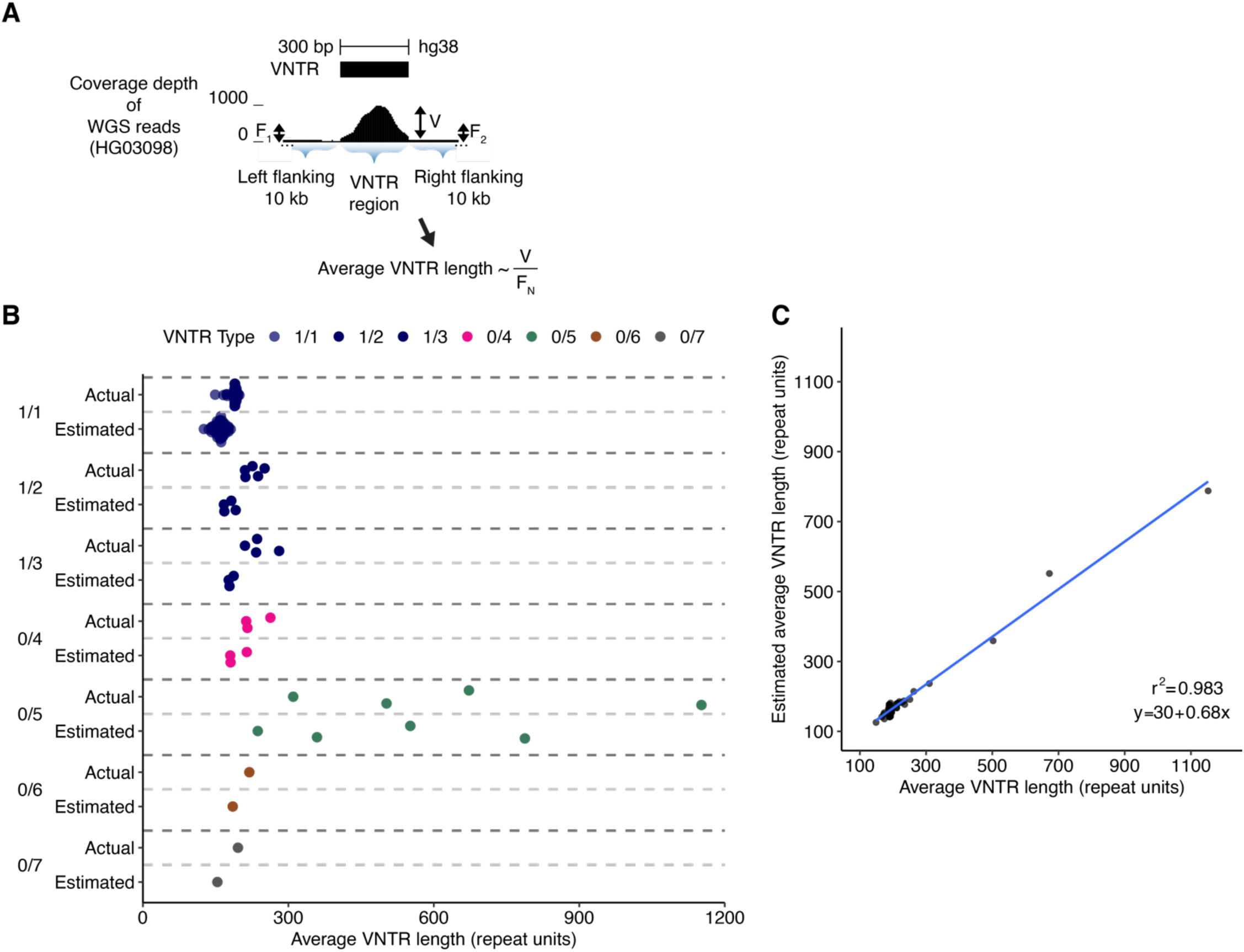
*CACNA1C* VNTR length estimates using short-read WGS data. (***A***) Schematic for estimation of *CACNA1C* VNTR length (in repeat units). Estimate corresponds to the average of both alleles within a diploid genome. Average sequencing coverage depth is computed across three regions: the *CACNA1C* VNTR region (V) and its two flanking 10-kb segments (F_1_ and F_2_). Average VNTR length (in repeat units) is computed as the average of V/F_1_ and V/F_2_ scaled by a conversion factor. (***B***) Average VNTR length is estimated for 70 individuals from short-read WGS data from 1000 Genomes Project 30X on GRCh38. Matching long-read assemblies are used to establish actual length; length is averaged across both alleles from the same individual. (***C***) Scatterplot of VNTR lengths estimated from WGS (averaged across each individual’s two alleles, y-axis) vs. actual lengths measured directly from long-read haplotype assemblies (n = 140 VNTR sequences, n = 70 diploid individuals).

## SUPPLEMENTARY TABLES

**Table S1.** Summary of long-read haplotype assemblies by project and read type.

| Project (Read Type) | Num. assemblies | Num. individuals |
| --- | --- | --- |
| HGSVC2 (CLR) | 42 | 21 |
| HGSVC2 (HiFi) | 26 | 13 |
| HPRC (HiFi) | 86 | 43 |
| CHM13 (HiFi) | 1 | 1 |
| Total | 155 | 78 |

Num. assemblies, count of long-read haplotype assemblies. Num. individuals, number of individual humans.

**Table S2.** Count of haplotype assemblies for each Type by long-read technology.

|  | PacBio HiFi | PacBio CLR |
| --- | --- | --- |
| Type 1 | 96 | 38 |
| Type 2 | 6 | 2 |
| Type 3 | 2 | 2 |
| Type 4 | 3 | 0 |
| Type 5 | 4 | 0 |
| Type 6 | 1 | 0 |
| Type 7 | 1 | 0 |
| Total | 113 | 42 |

**Table S3.** Nucleotide differences among VR2 alleles.

|  | VR2A | VR2B | VR2C |
| --- | --- | --- | --- |
| VR2B | 257 | – | – |
| VR2C | 226 | 44 | – |
| VR2D | 675 | 434 | 468 |

Number of mismatched nucleotide positions (including gaps) between each pair of aligned VR2 consensus sequences.

**Table S4.** Nucleotide differences between consensus and non-consensus VR alleles.

|  | A | B | C | D |
| --- | --- | --- | --- | --- |
| VR1 | 25 | 135 | – | – |
| VR2 | 33 | 46 | 35 | 40 |

Average number of mismatched nucleotide positions (including gaps) between consensus and non-consensus sequences of a VR allele. There are no VR1C or VR1D alleles.

**Table S5.** Association of VR1 and VR2 in Type 1 sequences.

|  | VR2A | VR2B | VR2C | VR2D | Unclassified |
| --- | --- | --- | --- | --- | --- |
| VR1A | 66 | 2 | 11 | 7 | – |
| VR1B | 1 | 45 | 0 | 0 | – |
| Unclassified | – | – | – | – | 2 |

Number of Type 1 sequences (n = 134) with each combination of VR1 and VR2 alleles.

**Table S6.**
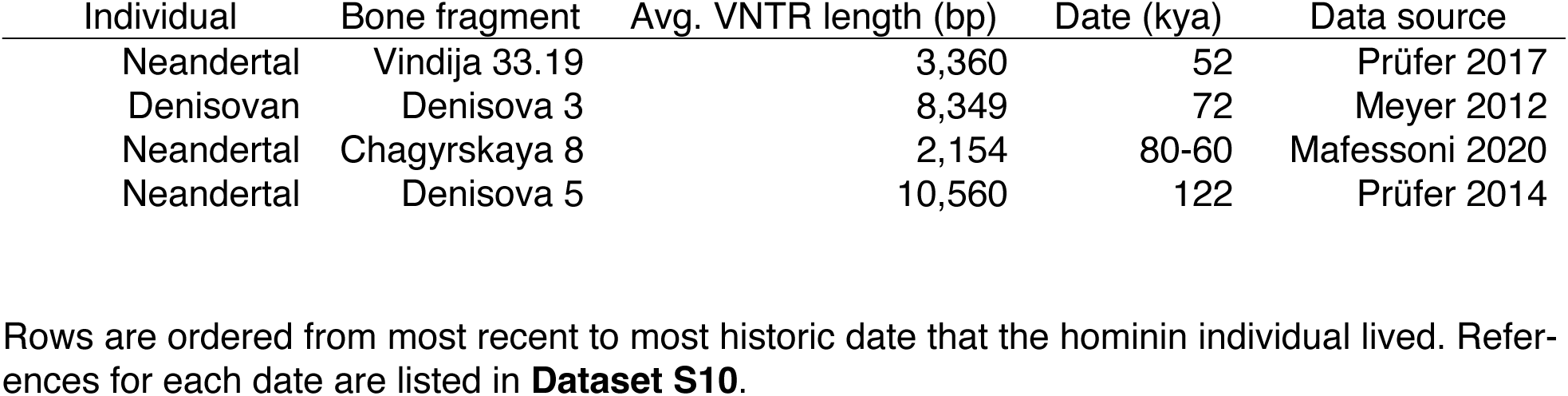
Estimated average *CACNA1C* VNTR length in four ancient hominin genomes.

## SUPPLEMENTARY DATA

### Dataset S1. Data sources for long-read genome assemblies and short-read WGS

Individual, the seven-character ID assigned to each individual by 1000 Genomes, Genome in a Bottle (NA24385 (HG002) and NA24631 (HG005)), or HapMap (NA12878 (HG001)). Project, the creator of the assembly. Read Type, PacBio long-read technology used to create initial phased contig assemblies. HiFi or CLR Coverage, reported as data yield of PacBio reads (Gbp) divided by estimated genome size (for HGSVC2 and HPRC individuals). For T2T, HiFi coverage is reported as the mean coverage of all chromosomes. Values compiled from the supplementary information of the relevant publications: (52) Table S6, (54) Table S1, (53) main text. Assembly 1 ID and Assembly 2 ID, a unique name for each haplotype assembly shortened from their filename. Assembly 1 Download Path and Assembly 2 Download Path, link to download each assembly in FASTA format. WGS Download Path, link to download each individual’s high-coverage WGS data from 1000 Genomes, when available. 1000 Genomes Population Code, population membership of each individual when available, as defined by (90).

### Dataset S2. Alignment of all *CACNA1C* VNTR sequences

All *CACNA1C* VNTR sequences analyzed in this manuscript, aligned within each Type and provided in FASTA format. Sequences in this file are ordered numerically by Type then alphabetically by assembly ID (**Dataset S1**).

### Dataset S3. Coordinates of VNTR in each assembly

Assembly ID, individual and haplotype. Filename, name of FASTA file containing genomic assembly. Region (contig:start-end), coordinates of *CACNA1C* VNTR in each assembly.

### Dataset S4. Repeat unit counts

Repeat unit, the 158 unique repeat units identified from the *CACNA1C* VNTR sequences analyzed in this manuscript. Length, length in nucleotides of each repeat unit. Count, number of occurrences of each repeat unit in 155 *CACNA1C* VNTR sequences. Frequency, global frequency of each repeat unit. Assembly IDs, assembly IDs of *CACNA1C* VNTR sequences containing at least 1 copy of a repeat unit. Num. assemblies, count of assemblies. Maximum value is 155, inclusive of *CACNA1C* VNTR sequences from the selected HGSVC2, HPRC, and T2T assemblies. Frac. assemblies, fraction of 155 assemblies containing at least 1 copy of a repeat unit.

### Dataset S5. Samples for validation of *CACNA1C* VNTR assembly

Assembly ID, individual and haplotype. Type, at least one sequence per Type is included. VR Alleles, Variable Region allele(s) and copy number. Project, the creator of the assembly. Read Type, PacBio long-read technology used to create initial phased contig assemblies. PacBio HiFi Reads Source Directory, link to directory with whole-genome BAM files of PacBio HiFi reads (or FASTQ files in the case of the HGSVC2-sourced sample HG03486_h2).

### Dataset S6. *CACNA1C* VNTR Type consensus sequences

Consensus sequences for *CACNA1C* VNTR Types described in this manuscript, provided in FASTA format.

Consensus sequences for each Type were converted to nucleotide sequences by swapping each character for its corresponding repeat unit. Types 2, 4, and 5 had characters representing infrequent 30-bp repeat units in their consensus sequence at 1, 5, and 97 positions, respectively. These characters were converted to the most common infrequent unit at each position. Most of these positions (75%, 77/103) contained the same infrequent unit that was not an alignment gap in all sequences. One position in Type 5 had two infrequent units in equal proportion. In this case, the repeat unit with a higher global frequency was chosen for the consensus. The remaining positions (24%, 25/103) had one infrequent unit that was not an alignment gap in a majority of sequences. Only one repeat unit in any consensus sequence was a repeat unit of size other than 30 bp. It was shared by 3/4 *CACNA1C* VNTR sequences in Type 5.

### Dataset S7. Duplications in Type 1 sequences

Alignment positions (repeat units), range of repeat unit positions in the aligned Type 1 consensus sequence corresponding to a tandemly duplicated sequence. Length (repeat units), length of duplicated sequence in repeat units. Length (nucleotides), length of duplicated sequence in nucleotides. Num. mismatches (nucleotides), number of nucleotide mismatches between a duplication source and destination sequence. Freq., frequency of duplicated sequence in 134 Type 1 *CACNA1C* VNTR sequences. Assembly IDs, assembly IDs of *CACNA1C* VNTR sequences with each duplication. Num. assemblies, count of assemblies with each duplication. Ancestries, ancestries of assemblies with each duplication. Duplication sequence, nucleotide sequence of the duplication. Mean repeat unit freq., average repeat unit frequency of those overlapping each gap in the Type 1 consensus. Min repeat unit freq., minimum repeat unit frequency of those overlapping each gap in the Type 1 consensus. Max repeat unit freq., maximum repeat unit frequency of those overlapping each gap in the Type 1 consensus.

### Dataset S8. VR1 alleles

Detail of VR1 alleles shown in **Fig. S8**. Allele, a unique identifier for each distinct VR1 sequence. Consensus alleles do not have a number appended to their identifier. Count, count of each VR1 sequence found in Type 1, 2, and 3 *CACNA1C* VNTR sequences. Duplicated VR1 sequences are counted according to their copy number. Assembly IDs, assembly IDs of *CACNA1C* VNTR sequences with each VR1 sequence. Assemblies with a duplicated VR1 sequence have each copy indicated by an appended integer. Num. mismatches, number of repeat unit mismatches between each VR1 sequence and the consensus sequence of its VR1 allele. The unclassified VR1 sequence has 10 and 9 mismatches (in repeat units) to VR1A and VR1B, respectively. Sequence, VR1 nucleotide sequences aligned to each other. Type, the *CACNA1C* VNTR Types each VR1 sequence is found in.

### Dataset S9. VR2 alleles

Detail of VR2 alleles shown in **Fig. S9**. Allele, a unique identifier for each distinct VR2 sequence. Consensus alleles do not have a number appended to their identifier. Count, count of each VR2 sequence found in Type 1, 2, and 3 *CACNA1C* VNTR sequences. Assembly IDs, assembly IDs of *CACNA1C* VNTR sequences with each VR2 sequence. Num. mismatches, number of repeat unit mismatches between each VR2 sequence and the consensus sequence of its VR2 allele. The unclassified VR2 sequence found in Type 1 (Unclassified-1) has 12, 20, 21, and 30 mismatches (in repeat units) to VR2A, VR2B, VR2C, and VR2D, respectively. The unclassified VR2 sequence found in Type 2 (Unclassified-2) has 23, 30, 29, and 15 mismatches (in repeat units) to VR2A, VR2B, VR2C, and VR2D, respectively. Sequence, VR2 nucleotide sequences aligned to each other. Type, the *CACNA1C* VNTR Types each VR2 sequence is found in.

### Dataset S10. References for divergence events mentioned in this paper

Event, historical event. Date, estimated date of historical event in thousand years ago (kya) or million years ago (mya). Publication, paper reporting the estimated date. Reference, numbered reference in this paper. Rows in this Dataset are ordered chronologically from most historic to most recent.

